# Poor neutralization and rapid decay of antibodies to SARS-CoV-2 variants in vaccinated dialysis patients

**DOI:** 10.1101/2021.10.05.21264054

**Authors:** Jessica Bassi, Olivier Giannini, Chiara Silacci-Fregni, Laura Pertusini, Paolo Hitz, Tatiana Terrot, Yves Franzosi, Francesco Muoio, Christian Saliba, Marcel Meury, Exequiel A. Dellota, Josh Dillen, Patrick Hernandez, Nadine Czudnochowski, Elisabetta Cameroni, Nicola Beria, Mariangela Ventresca, Alberto Badellino, Soraya Lavorato-Hadjeres, Elisabetta Lecchi, Tecla Bonora, Matteo Mattiolo, Guido Trinci, Daniela Garzoni, Giuseppe Bonforte, Valentina Forni-Ogna, Davide Giunzioni, Lorenzo Berwert, Ravindra K. Gupta, Paolo Ferrari, Alessandro Ceschi, Pietro Cippà, Davide Corti, Antonio Lanzavecchia, Luca Piccoli

**Affiliations:** Humabs Biomed SA, a subsidiary of Vir Biotechnology, Bellinzona, Switzerland; Faculty of Biomedical Sciences, Università della Svizzera italiana, Lugano, Switzerland; Department of Medicine, Ente Ospedaliero Cantonale, Bellinzona, Switzerland; Division of Nephrology, Ente Ospedaliero Cantonale, Lugano, Switzerland; Clinical Trial Unit, Ente Ospedaliero Cantonale, Lugano, Switzerland; Vir Biotechnology, San Francisco, California, United States of America; Cambridge Institute of Therapeutic Immunology & Infectious Disease (CITIID), Cambridge, UK; Department of Medicine, University of Cambridge, Cambridge, United Kingdom; Clinical School, University of New South Wales, Sydney, Australia; Division of Clinical Pharmacology and Toxicology, Institute of Pharmacological Science of Southern Switzerland, Ente Ospedaliero Cantonale, Lugano, Switzerland; Department of Clinical Pharmacology and Toxicology, University Hospital Zurich, Zurich, Switzerland; Faculty of Medicine, University of Zurich, Zurich, Switzerland

## Abstract

Patients on dialysis are at risk of severe course of SARS-CoV-2 infection. Understanding the neutralizing activity and coverage of SARS-CoV-2 variants of vaccine-elicited antibodies is required to guide prophylactic and therapeutic COVID-19 interventions in this frail population. By analyzing plasma samples from 130 hemodialysis and 13 peritoneal dialysis patients after two doses of BNT162b2 or mRNA-1273 vaccines, we found that 35% of the patients had low-level or undetectable IgG antibodies to SARS-CoV-2 Spike (S). Neutralizing antibodies against the vaccine-matched SARS-CoV-2 and Delta variant were low or undetectable in 49% and 77% of patients, respectively, and were further reduced against other emerging variants. The fraction of non-responding patients was higher in SARS-CoV-2-naïve hemodialysis patients immunized with BNT162b2 (66%) than those immunized with mRNA-1273 (23%). The reduced neutralizing activity correlated with low antibody avidity. Patients followed up to 7 months after vaccination showed a rapid decay of the antibody response with an average 21- and 10-fold reduction of neutralizing antibodies to vaccine-matched SARS-CoV-2 and Delta variant, which increased the fraction of non-responders to 84% and 90%, respectively. These data indicate that dialysis patients should be prioritized for additional vaccination boosts. Nevertheless, their antibody response to SARS-CoV-2 must be continuously monitored to adopt the best prophylactic and therapeutic strategy.

## Introduction

Patients with chronic kidney disease (CKD), in particular those with end-stage kidney disease (ESKD) on dialysis, are highly predisposed to infections, which are the major cause of morbidity and the second cause of mortality in this vulnerable population[1, 2]. Infection by SARS-CoV-2, the causative agent of the Coronavirus disease-2019 (COVID-19), has posed a new threat for CKD patients, who were found to have a greater risk of severe COVID-19 course[3, 4]. Early reports indicated a case fatality ranging from 10 to 30% in patients on hemodialysis (HD)[5-10].

The rapid development of COVID-19 vaccines has provided an important strategy to prevent SARS-CoV-2 infection and, especially, severe COVID-19 course. In particular, mRNA-based vaccines developed by Pfizer/BioNTech (BNT-162b2) and Moderna (mRNA-1273) have demonstrated high safety and efficacy in healthy and at-risk individuals, including patients with chronic diseases, cancer and solid organ transplantation[11-14]. However, immunosuppressed patients, in particular those with hematological malignancies, autoimmune diseases and solid organ transplantations, were shown to mount a low antibody response to these vaccines[15-20]. Because of their immunological frailty, patients on dialysis were prioritized in international COVID-19 vaccination programs[21]. Recent studies in HD patients showed a delayed and lower serological response to vaccines and a rapid decline of anti-SARS-CoV-2 antibodies[19, 22-24]. These findings suggest an overall diminished vaccine response to SARS-CoV-2 in ESKD patients that is reminiscent of the low response observed after vaccination against Hepatitis B virus (HBV) and seasonal Influenza virus[25, 26]. In addition, the rapid increase in cases of infection by SARS-CoV-2 Delta variant (B.1.617.2 lineage) since April 2021 provided a new potential challenge for dialysis patients, especially those with a suboptimal response to the vaccine[27].

At this stage of the pandemic, it is urgent to identify populations not developing sufficient levels of neutralizing antibodies against circulating SARS-CoV-2 variants, which therefore may be at-risk of developing severe COVID-19. In this study, we provide evidence of a poor neutralizing and rapidly decaying antibody response to mRNA-vaccines in the dialysis population, which supports the strategy of prioritizing these patients for an additional boost and other therapeutic strategies.

## Material and methods

### Study participants and ethics statement

Blood samples were obtained from 143 dialysis patients and 48 healthcare workers under study protocols approved by the local Institutional Review Board (Canton Ticino Ethics Committee, Switzerland). All the participants were recruited from the four public hospitals of the Ente Ospedaliero Cantonale (EOC) in Ticino (Southern Switzerland). All subjects provided written informed consent for the use of blood and blood components (such as PBMCs, sera or plasma).

### Isolation of plasma

Blood samples were collected from all the participants before, 2-3 weeks and up to 7 months after the second dose. An additional sample was collected 2-3 weeks after the first COVID-19 vaccine dose from 26 patients and all the 48 healthcare workers. Plasma was isolated from blood draw performed using BD tubes containing Ficoll (BD, CPT Ficoll, Cat. No. 362780) and stored at +4°C until use.

### Cell lines

Cell lines used in this study were obtained from ATCC (Vero E6 TMPRSS2) or ThermoFisher Scientific (Expi CHO cells, Expi293F™ and HEK293F cells). Vero E6 TMPRSS2 cells were grown in DMEM supplemented with 10% HyClone (FBS). Expi293F cells were grown in Expi293 Expression Medium.

### Production of recombinant glycoproteins

The SARS-CoV-2 RBD WT construct was synthesized by GenScript into phCMV1, with a sequence encoding an N-terminal mu-phosphatase signal peptide, an ‘ETGT’ linker, SARS-CoV-2 S residues 328-531, a linker sequence, an Avi tag, a twin Strep tag and a 8xHis-tag. Recombinant ACE2 (UniProt Q9BYF1, residues 19-615 with a C-terminal thrombin cleavage site-TwinStrep-10xHis-GGG-tag, and N-terminal signal peptide) and RBD WT constructs were transiently transfected into Expi293F cells following manufacturer’s instructions as previously described. Supernatants were clarified by centrifugation and affinity purified using a 5 mL StrepTrap column (28-9075-48, VWR). The SARS-CoV-2 stabilized Spike WT (D614G) construct was synthesized by GenScript into pCDNA3.1 with an N-terminal mu-phosphatase signal peptide, 2P stabilizing mutation[28, 29], a TEV cleavage site and a C-terminal foldon, 8x His-tag, Avi tag and C-tag[30] and expressed in HEK293F 293 cells following manufacturer’s instructions. Supernatants were clarified by centrifugation and affinity purified using a 5 mL C-tag affinity matrix column.

### VSV Spike mutants and pseudovirus generation

Amino acid substitutions were introduced into the D614G pCDNA_SARS-CoV-2_S plasmid as previously described[31]. SARS-CoV-2 S glycoprotein-encoding-plasmids used to produce SARS-CoV-2 VSV, namely the Wuhan prototype (D614, referred as WT) and variants Alpha (B.1.1.7), Beta (B.1.351), Gamma (P.1), Delta (B.1.617.2), Epsilon (B.1.429), Kappa (B.1.617.1) and Lambda (C.37), were obtained using a multistep based on overlap extension PCR (oePCR) protocol[32].

Replication defective VSV pseudovirus[33] expressing SARS-CoV-2 Spike proteins corresponding to the different VOC were generated as previously described[34].

### Plasma pseudovirus neutralization assay

Vero E6-TMPRSS2 were grown in DMEM supplemented with 10% FBS and seeded into white bottom 96 well plates (PerkinElmer, 6005688), as previously described[35]. Conditions were tested in duplicate wells in each plate and at least six wells per plate contained untreated infected cells (defining the 0% of neutralization, “MAX RLU” value) and infected cells in the presence of S2E12 and S2X259 mAbs at 50 µg/mL each (defining the 100% of neutralization, “MIN RLU” value). Average of Relative light units (RLUs) of untreated infected wells (MAX RLU_ave_) was subtracted by the average of MIN RLU (MIN RLU_ave_) and used to normalize percentage of neutralization of individual RLU values of experimental data according to the following formula: (1-(RLU_x_ - MIN RLU_ave_) / (MAX RLU_ave_ – MIN RLU_ave_)) x 100. Data were analyzed and visualized with Prism (Version 9.1.0). Each neutralization experiment included a technical duplicate. The loss or gain of neutralization potency across Spike variants was calculated by dividing the variant ID_50_ by the parental ID_50_.

### Enzyme-linked immunosorbent assay (ELISA)

Spectraplate-384 with high protein binding treatment (custom made from Perkin Elmer) were coated overnight at 4°C with 5 µg/mL RBD or 1 µg/mL SARS-CoV-2 S protein in PBS. The day after plates were washed and blocked with Blocker Casein in PBS (Thermo Fisher Scientific, 37528) supplemented with 0.05% Tween 20 (Sigma Aldrich), 1h RT. Serial dilutions of plasma samples were then added to plates for 1h RT. Alkaline Phosphatase-conjugated goat anti-human IgG, IgM or IgA (Southern Biotech) were added to plates and incubated for 1h RT. 4-NitroPhenyl Phosphate (pNPP, Sigma-Aldrich, N2765-100TAB) substrate was then added and plates were read after 1 h (IgG) or 2 h (IgA and IgM) at 405 nm with a BioTek plate reader. Data were plotted and analyzed with GraphPad Prism software (version 9.1.0). For chaotropic ELISA, after incubation with plasma, plates were washed and incubated with a 1 M solution of sodium thiocyanate (NaSCN, Sigma 251410) for 1h RT. Avidity Index was calculated as the ratio (%) of the ED_50_ in presence and the ED_50_ in absence of NaSCN.

### Blockade of RBD binding to human ACE2

Plasma were diluted in PBS and mixed with SARS-CoV-2 RBD mouse Fc-tagged antigen (Sino Biological, 40592-V05H, final concentration 20 ng/mL) and incubated for 30 min at 37°C. The percentage of inhibition was calculated as follows: (1−(OD sample−OD neg ctr)/(OD pos ctr−OD neg ctr)]) × 100.

### Statistical analysis

The study was designed to have 80% power to detect a minimum 25% difference in total incidence of cases with poor neutralizing antibody response (i.e., low or undetectable plasma antibody titers) or in average neutralizing titers between dialysis patients and healthy controls as well as within the HD subgroups. Comparisons of means between two groups of unpaired data were made with Mann-Whitney rank test. Comparisons of means between multiple groups of unpaired data were made with Kruskal-Wallis rank test and corrected with Dunn’s test. Comparisons of means between multiple groups of matched data were made with Friedman rank test and corrected with Dunn’s test. Relative risks of poor neutralizing response in selected groups of patients were calculated from 2×2 contingency tables using two-sided Fisher’s exact test. Statistical significance is set as P<0.05 and P-values are indicated with: ns=non-significant; *=0.033; **=0.002; ***<0.001. ED50 and ID50 titers were calculated from the interpolated value from the log(agonist) and the log(inhibitor), respectively, versus response, using variable slope (four parameters) non-linear regression. Data were plotted and analyzed with GraphPad Prism software (version 9.1.0).

## Results

### Naïve dialysis patients produce low or undetectable levels of antibodies after two vaccine doses

At the beginning of the COVID-19 vaccination campaign in January 2021, 143 dialysis patients (130 HD, of whom 68% on hemodiafiltration, and 13 on peritoneal dialysis, PD) were enrolled in this study from four dialysis units of the Ente Ospedaliero Cantonale in Ticino, Switzerland, and received two doses of SARS-CoV-2 mRNA vaccines (83% with BNT162b2 from Pfizer-BioNTech and 17% with mRNA-1273 from Moderna) between January and May 2021. Socio-demographic data, dialysis features, comorbidities, therapies and information about previous vaccinations and infections are summarized in **Table 1**. Of note, twenty-four (17%) dialysis patients were previously infected with SARS-CoV-2, of whom 18 (75%) had severe COVID-19 requiring hospitalization (13) or admission to an intensive care unit (5). A group of 48 healthcare workers, who received two doses of BNT162b2 vaccine between April and June 2021, were included as healthy controls (HC). Twenty-four HC were previously infected with SARS-CoV-2 reporting mild symptoms that did not require hospitalization.

**Table 1.** Socio-demographic and clinical data of enrolled dialysis patients and healthy controls.

|  | Dialysis patients | Hemodialysis (HD) | Peritoneal dialysis (PD) | Healthy controls (HC) |
| --- | --- | --- | --- | --- |
| Enrolled participants | 143 | 130 | 13 | 48 |
| Age, median years (min-max, IQR) | 76 (29-97, 67-83) | 76 (29-97, 68-83) | 71 (32-82, 55-78) | 42 (24-67, 32-49) |
| Gender, female, n (%) | 48 (34%) | 43 (33%) | 5 (38%) | 35 (73%) |
| BMI, median Kg/m<sup>2</sup> (min-max, IQR) | 25.9 (17.6-58.8, 22.9-29.2) | 26.0 (17.6-58.8, 22.8-29.4) | 25.7 (18-36.8, 23.3-29.0) | 25.0 (17-3, 21-28) |
| Smoker, n (%) | 21 (15%) | 18 (14%) | 3 (23%) | 13 (27%) |
| Dialysis treatment, n (%) |  |  |  |  |
| HD, n (%) | 130 (91%) | 130 (100%) |  |  |
| HDF, n (%) |  | 89 (68%) |  |  |
| PD, n (%) | 13 (9%) |  | 13 (100%) |  |
| Dialysis vintage, median months (min-max, IQR) | 42 (11-268, 25-80) | 46 (11-268, 26-87) | 28 (11-70, 16-44) |  |
| Dialysis access, native AVF, n (%) | 96 (67%) | 96 (74%) |  |  |
| Comorbidities, n (%) |  |  |  |  |
| Diabetes | 58 (41%) | 56 (43%) | 2 (15%) | 2 (4%) |
| Hypertension | 121 (85%) | 109 (84%) | 12 (92%) | 5 (10%) |
| Heart failure | 19 (13%) | 18 (14%) | 1 (8%) | - |
| Coronary heart disease | 46 (32%) | 41 (32%) | 5 (38%) | - |
| Peripheral arterial disease | 29 (20%) | 28 (22%) | 1 (8%) | - |
| Pulmonary disease | 42 (29%) | 38 (29%) | 4 (31%) | - |
| Chronic liver disease | 12 (8%) | 12 (9%) | - | - |
| Gastrointestinal disease | 27 (19%) | 24 (18%) | 3 (23%) | - |
| Hemato-oncological disease | 12 (8%) | 12 (9%) | 1 (8%) | - |
| Autoimmune disease | 11 (8%) | 9 (7%) | 2 (15%) | 1 (3%) |
| Kidney transplant | 10 (7%) | 10 (8%) | - | - |
| Neurodegenerative disease | 4 (3%) | 4 (3%) | - | - |
| Psychiatric disease | 13 (9%) | 13 (10%) | - | - |
| Charlson comorbidity index, median (min-max, IQR) | 8 (3-21, 7-11) | 9 (3-21, 7-11) | 6 (5-11, 6-8) | - |
| Previous SARS-CoV-2 infection<sup>1</sup>, n (%) | 24 (17%) | 23 (18%) | 1 (8%) | 24 (50%) |
| mild or no symptoms | 6 (25%) | 5 (22%) | 1 (100%) | 24 (50%) |
| severe course with hospitalization | 18 (75%) | 18 (78%) | - | - |
| ICU admission | 5 (28%) | 5 (28%) | - | - |
| Diagnosis-vaccine interval, median months (min-max, IQR) | 3.5 (1.1-9.7, 2.9-7.7) | 3.3 (1.1-9.7, 2.9-7.7) | 9.3 | 4.8 (3.5-12.8, 4.7-5.5) |
| COVID-19 vaccine received |  |  |  |  |
| Pfizer-BioNTech, n (%) | 118 (83%) | 107 (82%) | 11 (85%) | 48 (100%) |
| Moderna, n (%) | 25 (17%) | 23 (18%) | 2 (15%) | - |
| Dose 1 - dose 2 interval, median days (min-max, IQR) | 28 (20-38, 26-28) | 28 (20-28, 26-28) | 24 (21-28, 22-28) | 38 (23-31, 28-28) |
| Sampling after dose 1, median days (min-max, IQR) | 15 (9-33, 13-26) | 15 (9-33, 13-26) | - | 13 (12-16, 12-14) |
| Sampling after dose 2, median days (min-max, IQR) | 18 (13-27, 17-19) | 18 (13-27, 17-19) | 18 (16-25, 17-21) | 13 (10-21, 12-14) |
| 2020 seasonal flu vaccination, n (%) | 101 (70%) | 94 (72%) | 7 (54%) | 29 (60%) |
| Pharmacological treatment |  |  |  |  |
| Oral anticoagulation | 27 (19%) | 25 (19%) | 2 (15%) | - |
| Antiaggregant drugs | 124 (43%) | 124 (95%) | - | - |
| ACE inhibitors/ARBs | 70 (49%) | 58 (45%) | 12 (92%) | 3 (6%) |
| Calcium antagonists | 61 (43%) | 51 (39%) | 10 (77%) | - |
| Beta blockers | 88 (62%) | 79 (61%) | 9 (69%) | 2 (4%) |
| Statins | 80 (56%) | 71 (55%) | 9 (69%) | 2 (4%) |
| Oral antidiabetic drugs | 22 (15%) | 22 (17%) | - | 2 (4%) |
| Insulin | 37 (26%) | 35 (27%) | 2 (15%) | - |
| Immunosuppressive drugs<sup>2</sup> | 24 (17%) | 22 (17%) | 2 (15%) | - |
| Oral/intravenous steroids | 17 (12%) | 16 (12%) | 1 (8%) | - |
| Calcineurin inhibitors<sup>3</sup> | 8 (6%) | 8 (6%) | - | - |
| Vitamin D derivatives | 112 (78%) | 102 (78%) | 10 (77%) | - |

IQR, interquartile range. BMI, body mass index. HDF, hemodiafiltration. AVF, arteriovenous fistula. PCR, polymerase chain reaction. ICU, intensive care unit. ACE, angiotensin-converting enzyme. ARB, angiotensin receptor blockers. <sup>1</sup>with positive PCR and/or serology. <sup>2</sup>include any of steroids, cyclosporine, tacrolimus, rituximab, mycophenolate mofetil and azathioprine. <sup>3</sup>include cyclosporine and tacrolimus.

Plasma samples were collected 2 to 3 weeks after the first and the second dose and the levels of plasma IgG, IgA and IgM specific for SARS-CoV-2 Spike (S) were measured by ELISA. A single vaccine dose induced detectable IgG in 34.6% of dialysis patients compared to 89.6% of HC, with high antibody levels in all HD and HC participants, who had been previously infected with SARS-CoV-2, and in 29.2% of HD and 79.2% of HC, who had not been exposed to the virus (**S1 Fig**). Importantly, although seroconversion was observed in 94.4% of dialysis patients after the second vaccine dose, 35% of them had still low or undetectable levels of anti-SARS-CoV-2 S antibodies, compared to 100% of HC showing high levels of S-specific IgG (**Fig 1**a-b).

**Fig 1.**
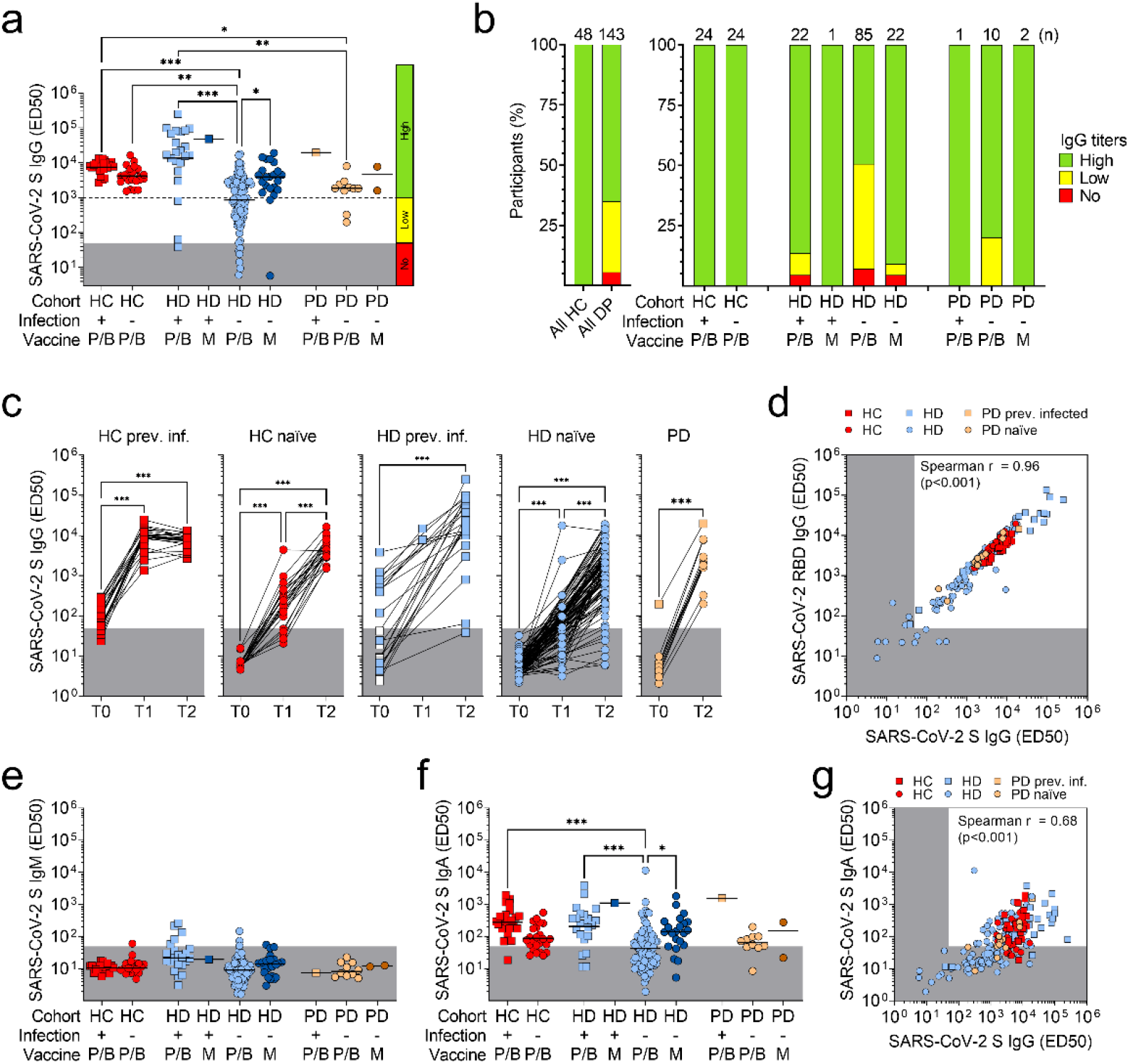
Comparison of mRNA vaccine-induced plasma antibody titers against SARS-CoV-2 between healthy controls and dialysis patients. a) Plasma IgG titers (ED50) to SARS-CoV-2 S after two doses of Pfizer/BioNTech (P/B) or Moderna (M) vaccines in previously infected (square) and naïve (circle) healthy controls (HC, red), hemodialysis (HD, blue) and peritoneal dialysis (PD, orange) patients. Grey areas indicate non-specific IgG titers <50, a cut-off that was determined on non-specific binding to uncoated ELISA plates. An additional cut-off of 1’000, determined from the lowest titers in HC after two doses, was used to distinguish low (50-1’000) from high (>1’000) IgG titers. Statistical significance is set as P<0.05 and P-values are indicated with asterisks (*=0.033; **=0.002; ***<0.001). b) Percentages of participants with high, low or no plasma SARS-CoV-2 S-specific IgG after two doses of mRNA-vaccine. Total number of participants within each cohort of HC and dialysis patients (DP) and within each subgroup is shown at the top of each bar. c) Kinetics of plasma IgG titers to SARS-CoV-2 S measured before vaccination (T0), after one (T1) or two (T2) vaccine doses. Each line connects samples from the same individual. White squares indicate HD patients who were not yet infected at T0 sampling. d) Correlation analysis between plasma IgG titers to SARS-CoV-2 S and RBD in all the plasma samples collected after the second vaccine dose. e) Plasma IgM titers to SARS-CoV-2 S after two vaccine doses in HC, HD and PD patients. Grey areas indicate non-specific IgM titers <50. f) Plasma IgA titers to SARS-CoV-2 S after two vaccine doses in HC, HD and PD patients. Grey areas indicate non-specific IgA titers <50. g) Correlation analysis between plasma IgG and IgA titers to SARS-CoV-2 S in all the plasma samples collected after the second vaccine dose.

The larger fraction of participants with low or no antibodies (36.9%) was represented by the HD group, with naïve patients immunized with BNT162b2 showing the lowest levels of plasma IgG (50.6%) compared to those who received mRNA-1273 (9.1%) (**Fig 1**a-b). The lower IgG level in naïve HD patients was the result of a slower kinetics of antibodies induced by the vaccines as compared to naïve HC and previously infected participants (**Fig 1**c). Of note, some previously infected HD patients produced higher amounts of antibodies compared to HC (**Fig 1**a), a finding that is consistent with a more severe COVID-19 course in these patients[36-38]. Plasma samples from the 13 PD patients were collected only after the second vaccine dose and all the participants showed detectable S-specific IgG with average plasma levels that were comparable to those of HC (**Fig 1**a-b). S-specific IgG levels highly correlated with RBD-specific IgG, suggesting that, similarly to natural infection, antibodies to this domain dominated the response induced by vaccination in both patients and controls (**Fig 1**d and **S2 Fig**, panel a). Finally, we observed that two vaccine doses induced, in the majority of the participants, undetectable levels of IgM and detectable levels of IgA specific for SARS-CoV-2 S, which were higher in previously infected HC and HD participants vaccinated with BNT162b2 and in naïve HD patients vaccinated with mRNA-1273 compared to naïve HD vaccinated with BNT162b2, and correlated with the corresponding IgG levels (**Fig 1**e-g and **S2 Fig**, panels b and c).

### Poor neutralization of Wuhan prototypic SARS-CoV-2 and related risk factors in naïve dialysis patients

To determine the neutralizing activity of vaccine-induced antibodies, we used plasma samples to perform an *in vitro* neutralization assay with pseudotyped vesicular stomatitis virus (VSV) that expresses the Wuhan wild-type (WT, D614) SARS-CoV-2 S glycoprotein (**Fig 2**a and **S3 Fig**, panel a). Compared to HC, 49% of dialysis patients had no or low neutralizing activity against the vaccine-matched SARS-CoV-2 strain (**Fig 2**b). In particular, while most of previously infected HD patients (87%) had moderate to high neutralizing antibody titers, which were similar or higher than those of HC, naïve HD patients had a heterogenous response with 57% of them characterized by a poor neutralizing activity, which was completely absent in the majority (60.7%) of these patients (**Fig 2**a). Of note, the fraction of non-responding patients was higher in naïve HD patients immunized with BNT162b2 (65.9%) than those immunized with mRNA-1273 (22.7%) (**Fig 2**b). Despite higher plasma S-specific IgG titers (**Fig 1**a-b), 60% of naïve PD patients had a similar poor neutralizing response compared to naïve HD patients, with 33.3% of those, who received BNT162b2 vaccine, showing no neutralizing activity (**Fig 2**a-b). The poor neutralizing activity was confirmed also by the low capability of plasma antibodies to inhibit binding of RBD to human ACE2 receptor (**S3 Fig**, panel b). In some previously infected dialysis patients, we observed neutralizing and ACE2-inhibiting antibody titers that were higher than those of HC and correlated with S- and RBD-specific IgG levels (**Fig 2**a and **S3 Fig**, panels c-e).

**Fig 2.**
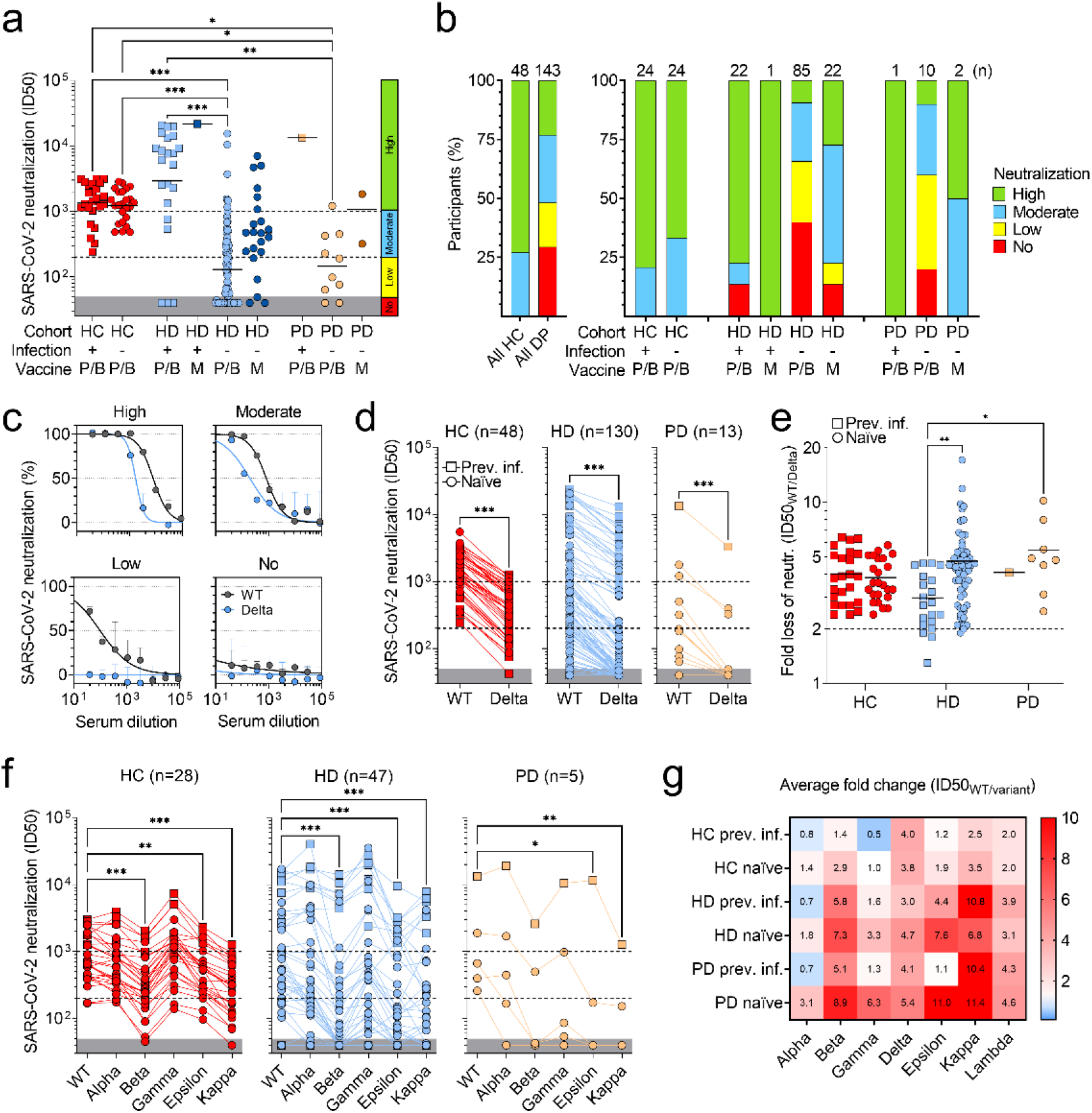
Analysis of neutralizing activity of plasma antibodies against wild-type SARS-CoV-2, Delta and other variants. a) Neutralizing antibody titers (ID50, 50% of inhibitory dilution) against pseudotyped VSV viruses harboring wild-type (D614) SARS-CoV-2 S determined using plasma from previously infected (square) and naïve (circle) healthy controls (HC), hemodialysis (HD) and peritoneal dialysis (PD) patients after two doses of Pfizer/BioNTech (P/B) or Moderna (M) vaccines. Grey areas indicate non-neutralizing titers (<50). A cut-off of 200, determined from the lowest neutralizing titers in HC, and a cut-off of 1’000, determined from the 25% percentile of titers in previously infected HC cohort, were used to distinguish low (50-200) from moderate (200-1’000) or high (>1’000) neutralizing titers. Statistical significance is set as P<0.05 and P-values are indicated with asterisks (*=0.033; **=0.002; ***<0.001). Shown are data from n=2 independent experiments. b) Percentages of participants with high, moderate, low or no plasma neutralizing antibodies to WT SARS-CoV-2 S after two doses of mRNA vaccine. Total number of participants within each cohort of HC and dialysis patients (DP) and within each subgroup is shown at the top of each bar. c) Neutralization of WT (black) and Delta (B.1.617.2, blue) SARS-CoV-2 pseudotyped VSV by four representative plasma samples showing high, moderate, low or no neutralization to WT SARS-CoV-2. d-e) Side-by-side comparison (d) and fold change analysis (e) of neutralizing titers against WT and Delta SARS-CoV-2 in 48 HC, 130 HD and 13 PD patients. Fold change is calculated as the ratio of ID50 value of WT and ID50 value of Delta variant. Shown are data from n=2 independent experiments. f) Side-by-side comparison of neutralizing titers against WT and Alpha (B.1.1.7), Beta (B.1.351), Gamma (P.1), Epsilon (B.1.429) and Kappa (B.1.617.1) SARS-CoV-2 variants in 28 HC, 47 HD and 5 PD patients. g) Fold change analysis of neutralizing titers against different variants. Numbers in each cell of panel g indicate the average fold change values of WT ID50 to variant ID50 in all the cohorts analyzed including only participants with ID50 neutralizing titers against WT SARS-CoV-2 greater than 80. Shown are data from n=2 independent experiments.

We next performed a sub-analysis to understand which socio-demographic and clinical data were associated to a significant risk for dialysis patients of being poor responders, here defined as having low or no neutralizing antibody titers after two mRNA-vaccine doses. Among all patients, dialysis mode (HD or PD) was not identified as risk factor, whereas vaccination with BNT162b2 and no previous SARS-CoV-2 infection represented the major risk factors for having a poor neutralizing antibody response (relative risk of 2.75 and 4.50, 95% confidence interval [CI] of 1.37 to 6.30 and 1.78 to 13.1, respectively) (**Table 2**). Within the larger naïve HD group, socio-demographic factors, including gender, age, body mass index, smoke and dialysis features could not be identified as risk factors (**Table 3**). Among comorbidities, we found that patients with heart failure history had an 1.68-increased relative risk of poor response (95% CI, 1.16 to 2.17). As expected, patients in therapy with immunosuppressive drugs had a 1.53 relative risk of poor response to the vaccine (95% CI, 1.05 to 2.02) with, in particular, calcineurin inhibitors accounting for the highest relative risk (1.87; 95% CI, 1.18 to 2.29). Vaccination with BNT162b2 was confirmed as a major factor with a relative risk of poor response of 2.88 (95% CI, 1.47 to 6.55), which increased to 3.60 (95% CI, 1.79 to 8.23) in 22 naïve HD patients who were matched to 22 naïve HD patients immunized with mRNA-1273 by age, gender, dialysis vintage and comorbidities. Among HD patients immunized with BNT162b2, those older than 80 and with heart failure history had an increased risk of poor response of 1.57 (95% CI, 1.18 to 2.08) and 1.55 (95% CI, 1.11 to 1.96), respectively (**Table 3**).

**Table 2.**
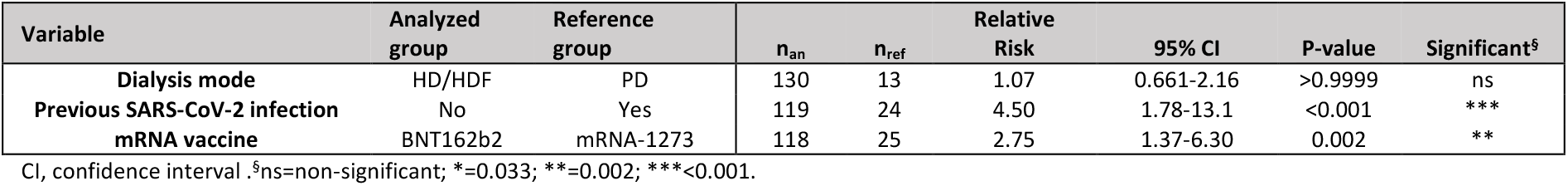
Analysis of relative risk of poor neutralizing response to COVID-19 vaccination in dialysis patients.

**Table 3.**
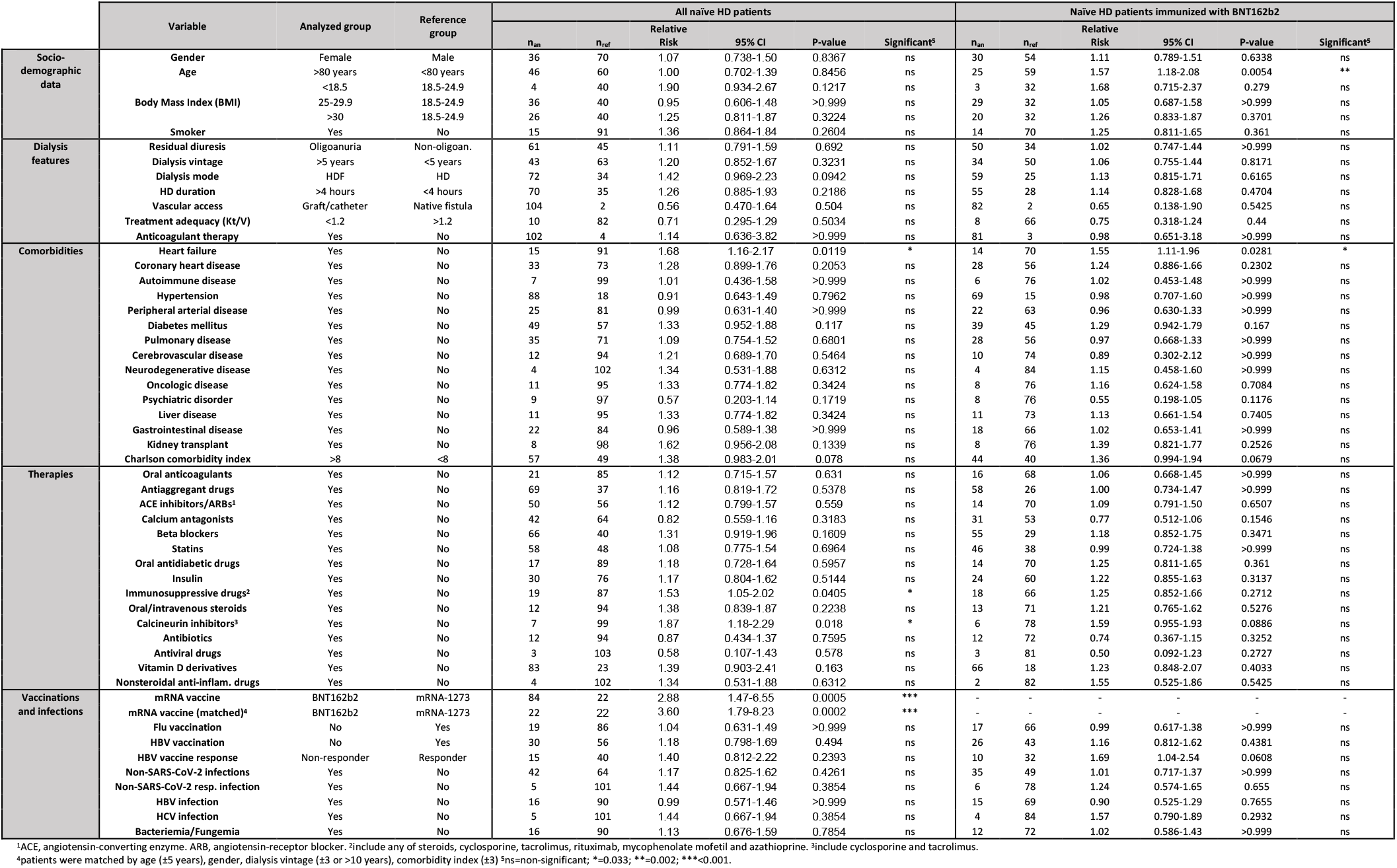
Analysis of relative risk of poor neutralizing response to COVID-19 vaccination in naïve HD patients.

### Loss of neutralization of Delta and other SARS-CoV-2 variants in dialysis patients

We next addressed the question whether vaccine-induced antibodies that neutralize wild-type SARS-CoV-2 could also neutralize current circulating SARS-CoV-2 variants, in particular the Delta variant (B.1.617.2 lineage) (**Fig 2**c). Most dialysis patients (76.9%) had plasma antibodies with low or undetectable neutralizing activity against the Delta variant, with 48.0% and 80.0% of naïve HD and PD patients with detectable levels of neutralizing antibodies against WT SARS-CoV-2 showing complete loss of neutralization against this variant (**Fig 2**d and **S4 Fig**, panel a). Almost all the participants, including HC, had a greater than 2-fold reduction in the neutralizing activity against Delta variant, with naïve HD patients showing up to 17-fold (average 4.7) loss in neutralization, as compared to previously infected patients that showed a maximum 5-fold loss (average 3.0) (**Fig 2**e). Similarly, naïve PD patients showed up to 10-fold (average 5.4) loss of neutralization against Delta variant (**Fig 2**e).

The loss of neutralization against some of the other circulating SARS-CoV-2 variants was higher compared to Delta, in particular for naïve HD patients, who showed an average 7.3-, 7.6- and 6.8-fold reduction in their neutralizing activity against Beta (B.1.351), Epsilon (B.1.429) and Kappa (B.1.617.1) variants (**Fig 2**f-g and **S4 Fig**, panels b-d). Neutralization of Alpha (B.1.1.7), Gamma (P.1) and Lambda (C.37) was also reduced in HD patients, but with minor loss compared to Delta (1.8-, 3.3- and 3.1-fold reduction, respectively) (**Fig 2**f-g and **S4 Fig**, panels e-h). Previously infected HD patients and naïve PD patients also showed a drastic loss of neutralization against different variants, which was comparable to that of naïve HD patients, but higher than that of HC (**Fig 2**f-g and **S4 Fig**). Complete loss of neutralization of Beta, Epsilon and Kappa variants was observed in 37-43.3% of HD and 75% of PD patients with detectable neutralizing antibodies against WT SARS-CoV-2 (**S4 Fig**, panel a).

### The reduced neutralizing activity correlates with a low antibody avidity

To further characterize the poor neutralizing activity of vaccine-induced antibodies, we determined the avidity of plasma antibodies by measuring their binding to SARS-CoV-2 S in presence of sodium thiocyanate, a chaotropic agent that induces dissociation of the antibody from the antigen in case of low affinity. While most of the previously infected HD patients (87%) showed moderate to high-avidity antibodies, a high fraction of naïve HD showed low-(42%) or no (9%) avidity antibodies and most of the naïve PD patients (75%) also showed low-avidity antibodies (**Fig 3**a). As expected, antibodies of all HC that were previously infected with SARS-CoV-2 showed high avidity (index >50%), whereas naïve HC showed moderate avidity (23-50%), consistent with the different duration of affinity maturation ongoing in the two groups (**Fig 3**a). The avidity indexes correlated with SARS-CoV-2 S-specific antibody and neutralizing titers (**Fig 3**b and **S5 Fig**, panel a). In particular, we observed larger fractions of HD patients with moderate to high avidity among patients with higher neutralizing antibody titers (**Fig 3**c). The lower avidity titers were paralleled by higher fractions of dialysis patients showing low- or non-neutralizing antibodies against the Delta and other SARS-CoV-2 variants (**Fig 3**d and **S5 Fig**, panels b-f).

**Fig 3.**
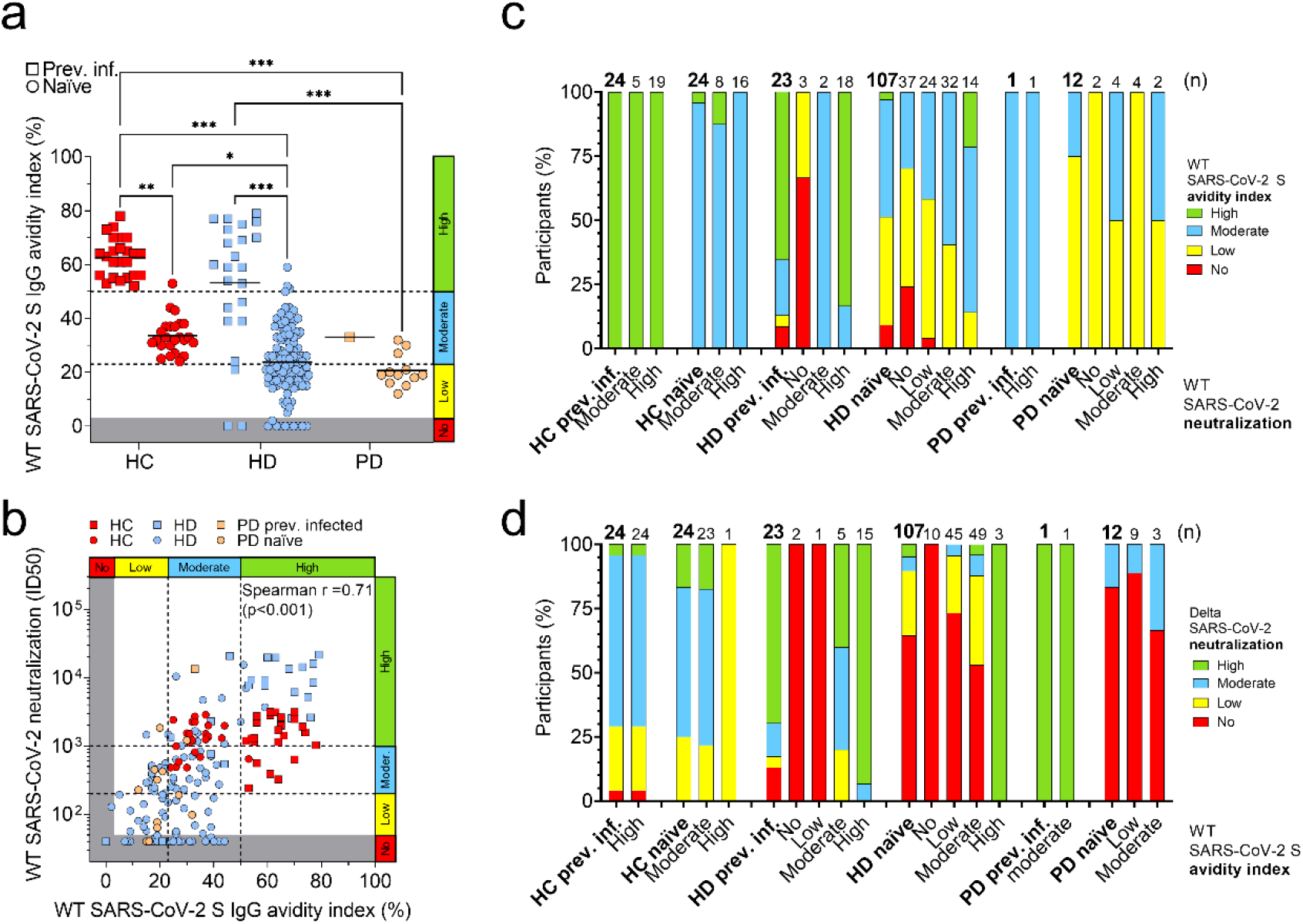
Role of avidity of plasma antibodies in the neutralization of wild-type SARS-CoV-2 and Delta variant. a) Avidity index of plasma antibodies to WT SARS-CoV-2 S in previously infected (square) and naïve (circle) healthy controls (HC), hemodialysis (HD) and peritoneal dialysis (PD) patients after two doses of mRNA vaccine. Grey areas indicate no avidity measured of plasma antibodies with ED50 titers lower than 50. A cut-off of 23%, determined from the lowest avidity index in naïve HC, and a cut-off of 50%, determined from the lowest avidity index in previously infected HC cohort, were used to distinguish low (0-23%) from moderate (23-50%) or high (>50%) avidity. Statistical significance is set as P<0.05 and P-values are indicated with asterisks (*=0.033; **=0.002; ***<0.001). Shown are data from n=2 independent experiments. b) Correlation analysis between plasma IgG avidity index to WT SARS-CoV-2 S and neutralization of WT SARS-CoV-2 in all the plasma samples collected after the second vaccine dose. c) Percentages of participants having plasma antibodies with high, moderate, low or no avidity to WT SARS-CoV-2 S after two doses of mRNA vaccine. Participants are shown as a total (bold) or divided by level of neutralization of WT SARS-CoV-2 (no, low, moderate, high). Total number of participants within each group is shown at the top of each bar. d) Percentages of participants having plasma antibodies with high, moderate, low or no neutralization of Delta SARS-CoV-2 after two doses of mRNA vaccine. Participants are shown as a total (bold) or divided by level of avidity to WT SARS-CoV-2 S (no, low, moderate, high). Total number of participants within each group is shown at the top of each bar.

### Rapid decay of neutralizing antibody in 7 months after vaccination

We followed 47 HC and 133 dialysis patients (121 HD and 12 PD) up to 7 months after vaccination (mean 5 months) and measured plasma titers and avidity of antibodies to SARS-CoV-2 S and their neutralizing activity against the vaccine-matched SARS-CoV-2 and Delta variant (**Table 4**). We found that 12.8% and 77.4% of HC and dialysis patients had low or undetectable S-specific IgG with a 4.2- and 5.2 fold-average reduction of IgG titers compared to the peak of the antibody response after the second vaccine dose (**Fig 4**a and **S6 Fig**, panel a). This titer decay was paralleled by a 12.7- and 21-fold average reduction of neutralizing antibodies to wild-type SARS-CoV-2 in HC and dialysis patients, which resulted in 40% and 84% of non-responders, respectively (**Fig 4**c and **S6 Fig**, panel c). In the case of antibodies neutralizing the Delta variant, we observed a 6.2- and 9.8-average reduction in HC and dialysis patients, which increased the fraction of non-responders to 72% and 90%, respectively (**Fig 4**d and **S6 Fig**, panel d). In line with the analysis at 2 weeks after the second dose, the most affected patients were those in hemodialysis, naïve to SARS-CoV-2 and vaccinated with BNT-162b2, with 97.5% and 99% of non-responders to WT SARS-CoV-2 and Delta variant, respectively (**Table 4**). Interestingly, only 36.2% and 38.8% of HC and dialysis patients showed an increase in the avidity index of SARS-CoV-2 S IgG, which, however, maintained its correlation with the neutralizing activity (**Fig 4**b and **S6 Fig**, panels b and e). Importantly, despite the overall waning of the antibody response during the follow-up, no new cases of SARS-CoV-2 infection were diagnosed in both HC and dialysis patients after vaccination.

**Table 4.**
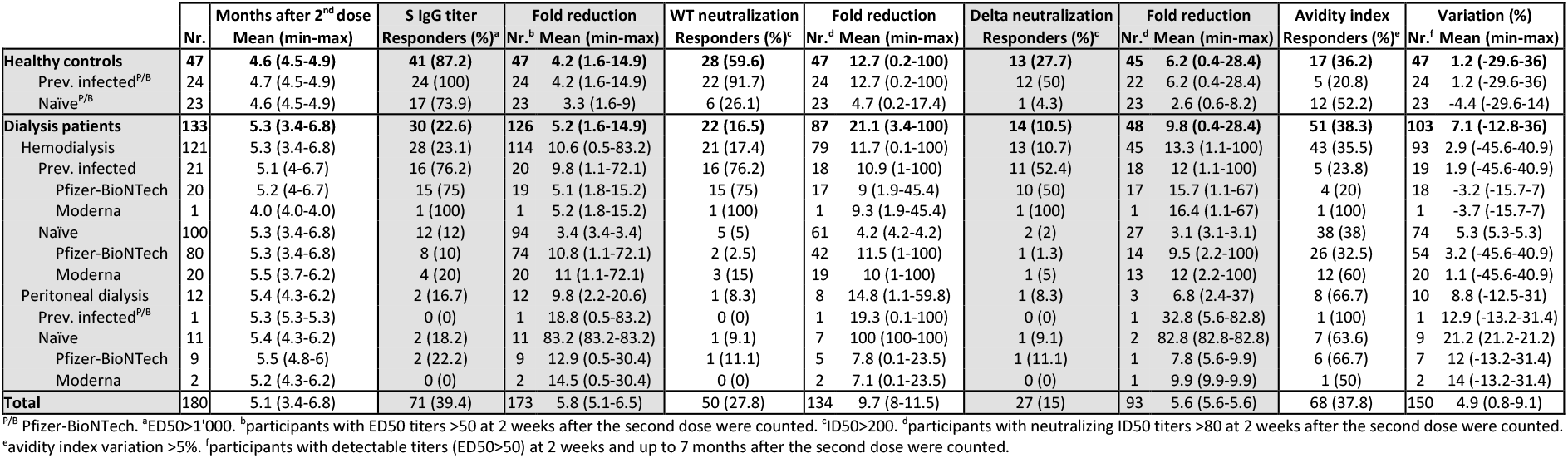
Follow-up analysis of SARS-CoV-2 S IgG titer, wild-type SARS-CoV-2 and Delta neutralization, and avidity index.

**Fig 4.**
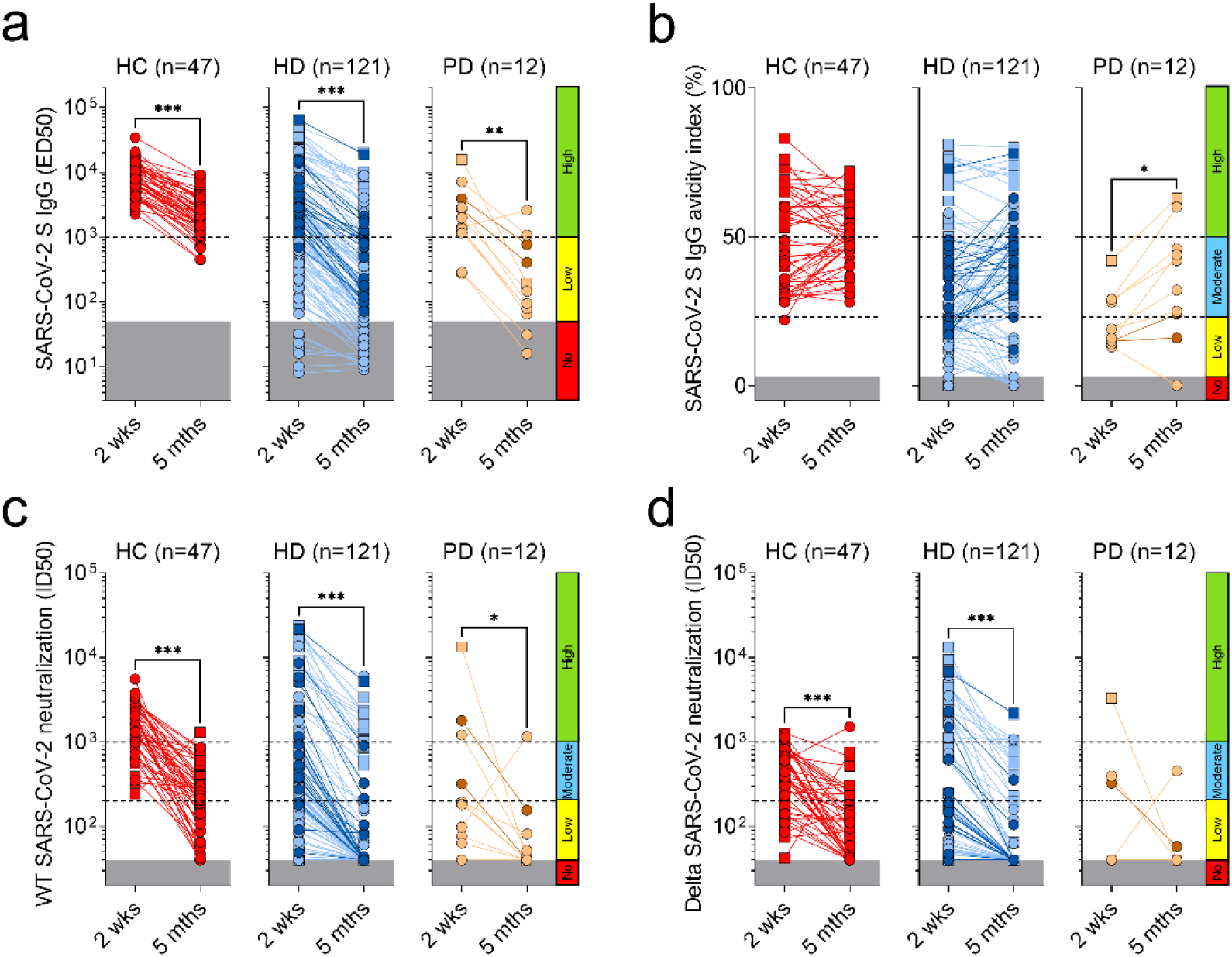
Follow-up analysis of SARS-CoV-2 S IgG, neutralizing titers and avidity index. a) Plasma IgG titers (ED50) to SARS-CoV-2 S after two doses of Pfizer/BioNTech (P/B, red, light blue and light orange) or Moderna (M, dark blue and dark orange) vaccines in samples collected from previously infected (square) and naïve (circle) healthy controls (HC, red), hemodialysis (HD, blue) and peritoneal dialysis (PD, orange) patients 2 weeks and up to 7 months (indicated mean 5 months) after vaccination. Cut-offs are shown as explained in Fig 1. Statistical significance is set as P<0.05 and P-values are indicated with asterisks (*=0.033; **=0.002; ***<0.001). b) Avidity index of plasma antibodies to WT SARS-CoV-2 S. Cut-offs are shown as explained in Fig 3. c, d) Neutralizing antibody titers (ID50) against pseudotyped VSV viruses harboring wild-type (D614) SARS-CoV-2 S (c) or Delta variant (d). Cut-offs are shown as explained in Fig 2.

## Discussion

In this phase of the COVID-19 pandemic where SARS-CoV-2 variants are rapidly spreading worldwide, we urgently need to understand the efficacy of vaccines administered so far in high-risk immunocompromised populations, in particular patients with chronic diseases[39, 40]. In this study, we provide evidence that dialysis patients do not develop sufficient levels of neutralizing antibodies against SARS-CoV-2 variants after two doses of mRNA vaccines.

The analysis of plasma SARS-CoV-2-specific IgG showed an overall response to mRNA vaccine in 94.4% dialysis patients, which is comparable to data shown in other reports (70.5-96%)[19, 22, 41, 42]. Nevertheless, we observed reduced antibody levels after both the first and the second dose in HD patients who were naïve to SARS-CoV-2, a finding consistent with a delayed immune response in this population[19, 43, 44]. These data suggest that measuring serum antibody titers to SARS-CoV-2 in dialysis patients could help clinicians to identify vaccine non-responders and guide clinical decision-making. However, currently available serological tests detect antibodies against the Spike of the Wuhan SARS-CoV-2 and do not define the level of neutralizing activity against the virus, which is considered a more relevant serological correlate of protection[45].

In this study, we found that 49% of dialysis patients had low or undetectable levels of neutralizing antibodies against the vaccine-matched SARS-CoV-2 with a significant reduction in neutralizing titers against different circulating variants in dialysis patients, which is consistent with an immune evasion by the virus[27, 35, 46]. Based on these data, we calculated that the fraction of dialysis patients non-responding to the vaccine further increased to 77% in the case of the Delta variant, suggesting that the great majority of the patients immunized with two doses of an mRNA vaccine may not develop a protective antibody response against SARS-CoV-2 variants. Importantly, follow-up data collected up to 7 months after vaccination showed a rapid decay of neutralizing antibodies to both wild-type SARS-CoV-2 and Delta variant, which increased the fraction of non-responders to 84% and 90%, respectively. These data suggest that dialysis patients have an increased risk of infection with possible severe outcomes over time from vaccination, an association which has been recently described in a US veteran cohort followed up to 6 months after vaccination[47]. However, we did not diagnose new cases of SARS-CoV-2 infection during the follow-up, which might be explained by the lowest incidence observed in Switzerland in the spring-summer season, and by the strict preventive measures adopted by the dialysis patients that might have compensated for the waning of the vaccine-induced antibodies.

Despite the huge discrepancy between the fraction of non-responders based on antibody levels and their neutralizing activity, we observed a high correlation between these titers, which may help to define proper cut-offs in the serological tests used in clinical settings to identify non-responding patients. Furthermore, the correlation observed between avidity and neutralization suggests that vaccine-induced antibodies require high avidity to neutralize SARS-CoV-2. Therefore, the identification of non-responders may be guided by a SARS-CoV-2 avidity test, as successfully implemented for other infectious diseases[48-52].

We identified that having never been exposed to SARS-CoV-2 is the major factor that increases the risk of a poor antibody response after vaccination, especially in HD patients. Indeed, naïve individuals were shown to develop lower titers of neutralizing antibodies to SARS-CoV-2 variants compared to previously infected individuals after two mRNA vaccine doses[35], indicating that vaccination may boost the neutralizing antibody response in dialysis patients after infection[53]. This effect can be explained by the higher antibody avidity that previously infected patients gained over time compared to naïve patients, who instead showed a delayed and low-avidity response. These findings are consistent with a slower affinity maturation of SARS-CoV-2 S-specific B cells, which appears to be a peculiar immunological feature of the dialysis population.

A recent report by Carr et al[54], showed that naïve HD patients vaccinated with BNT162b2 developed higher neutralizing antibodies against SARS-CoV-2 variants compared to Astra Zeneca - Oxford University AZD1222 vaccine, suggesting that mRNA vaccines provide a higher level of protection in dialysis patients compared to adenovirus-based vaccines. However, a difference in efficacy may exist among mRNA vaccines, as we found that being immunized with BNT162b2 instead of mRNA-1273 was the second most relevant risk factor for a poor response in the dialysis population, especially in naïve HD patients. This difference can be explained by the lower mRNA amount provided by one dose of BNT162b2 compared to mRNA-1273[11, 12]. Indeed, the mRNA-1273 vaccine was found to be more reactogenic[55] and to induce higher antibody titers in healthy individuals as well as in the more vulnerable elderly population[56-59].

Similarly to other studies analyzing the factors that were associated to low antibody levels after COVID-19 vaccination in dialysis patients, we also found a few risk factors of poor neutralizing antibody response to mRNA vaccines, including age, heart failure and immunosuppression[14, 20, 22, 23, 43, 60-62]. However, other factors, such as gender and dialysis vintage[43], dialysis adequacy measured by Kt/V[20], comorbidities[14] and non-responsiveness to Hepatitis B vaccination[20, 23] were not identified as risk factors in our study.

Our study has some limitations including a missing control group of patients matched by age, gender and comorbidities, without ESKD, while our control group was composed by younger individuals with few or no comorbidities. Another limitation was the low number of PD patients that did not allow to make subgroup analyses of risk factors of poor response. In addition, our patients were unbalanced in terms of type of mRNA vaccine received (BNT162b2 or mRNA-1273), although we had sufficient statistical power to make a comparison, even in a subgroup of patients matched by age, gender and comorbidity index.

In conclusion, our study demonstrates, at the functional level, that mRNA vaccines induce a poor neutralizing and rapidly decaying antibody response against SARS-CoV-2 variants in dialysis patients, in particular in naïve HD patients immunized with BNT162b2. Our findings support the need of an additional boost, preferentially with a high-dose mRNA vaccine, in this population[62-67], which, however, need to be continuously monitored with proper serological tests that measure not only the serum antibody levels, but also their neutralizing activity, either directly or indirectly through an avidity test. Finally, our data suggest that some patients may not respond efficiently even after an additional boost and, therefore, in case of SARS-CoV-2 infection, they should be considered for other therapeutic strategies, including early immunotherapy with monoclonal antibodies.

## Data Availability

All data produced in the present study are available upon reasonable request to the authors.

## Acknowledgments

We would like to thank all the nurses of the Ente Ospedaliero Cantonale, who helped collecting data and blood samples: Gianna Croppi, Sara Candolfi, Marie-Eve Brodeur, Chiara Tettamanti, Giorgia Esposito, Fabio Capocasale, Elisa Capitale, Marianna Vitini, Pedrag Lazarevic, Gaetano Donato, Fabienne Poncetta, Chiara Mattei. We thank Sarita Prosperi and Barbara Guggiari-De Rosa for organizational support. A special thanks to all the dialysis patients who accepted to participate to this study.

## Authors’ contributions

O.G., P.F., A.C., P.C., D.C, A.L. and L.Pi. contributed to study concept and design. L.Pe., N.B., A.B., S.L.-H., E.L., T.B., M.Ma., G.T., D.G., G.B., V.F.-O., D.G., L.B., A.R., contributed to donors’ recruitment, questionnaire and sample collection. L.Pe., T.T., Y.F., P.H. contributed to database preparation, data filling and cleaning. J.B., C.S.-F., F.M. contributed to sample processing. J.B., L.Pi. contributed to experiment design. J.B., C.S.-F., F.M. performed serological assays. M.Me., E.A.D.J., N.C., E.C. contributed to protein expression and purification. C.S., R.K.G. contributed to pseudotyped virus production. J.B., O.G., P.H., D.C., L.Pi. contributed to data analysis. J.B., O.G., P.F., A.C., P.C., D.C, A.L. and L.Pi. wrote the manuscript. O.G., L.Pi. provided overall supervision.

## References

1. Zoccali C, Vanholder R, Massy ZA, Ortiz A, Sarafidis P, Dekker FW, et al. The systemic nature of CKD. Nat Rev Nephrol. 2017;13(6):344–58. Epub 2017/04/25. doi: 10.1038/nrneph.2017.52. PubMed PMID: 28435157.

2. Dalrymple LS, Go AS. Epidemiology of acute infections among patients with chronic kidney disease. Clin J Am Soc Nephrol. 2008;3(5):1487–93. Epub 2008/07/25. doi: 10.2215/CJN.01290308. PubMed PMID: 18650409; PubMed Central PMCID: PMCPMC4571152.

3. Henry BM, Lippi G. Chronic kidney disease is associated with severe coronavirus disease 2019 (COVID-19) infection. Int Urol Nephrol. 2020;52(6):1193–4. Epub 2020/03/31. doi: 10.1007/s11255-020-02451-9. PubMed PMID: 32222883; PubMed Central PMCID: PMCPMC7103107.

4. Cippa PE, Cugnata F, Ferrari P, Brombin C, Ruinelli L, Bianchi G, et al. A data-driven approach to identify risk profiles and protective drugs in COVID-19. Proc Natl Acad Sci U S A. 2021;118(1). Epub 2020/12/12. doi: 10.1073/pnas.2016877118. PubMed PMID: 33303654; PubMed Central PMCID: PMCPMC7817222.

5. Anand S, Montez-Rath M, Han J, Bozeman J, Kerschmann R, Beyer P, et al. Prevalence of SARS-CoV-2 antibodies in a large nationwide sample of patients on dialysis in the USA: a cross-sectional study. Lancet. 2020;396(10259):1335–44. Epub 20200925. doi: 10.1016/S0140-6736(20)32009-2. PubMed PMID: 32987007; PubMed Central PMCID: PMCPMC7518804.

6. Corbett RW, Blakey S, Nitsch D, Loucaidou M, McLean A, Duncan N, et al. Epidemiology of COVID-19 in an Urban Dialysis Center. J Am Soc Nephrol. 2020;31(8):1815–23. Epub 2020/06/21. doi: 10.1681/ASN.2020040534. PubMed PMID: 32561681; PubMed Central PMCID: PMCPMC7460899.

7. Jager KJ, Kramer A, Chesnaye NC, Couchoud C, Sanchez-Alvarez JE, Garneata L, et al. Results from the ERA-EDTA Registry indicate a high mortality due to COVID-19 in dialysis patients and kidney transplant recipients across Europe. Kidney Int. 2020;98(6):1540–8. Epub 2020/09/27. doi: 10.1016/j.kint.2020.09.006. PubMed PMID: 32979369; PubMed Central PMCID: PMCPMC7560263.

8. Hilbrands LB, Duivenvoorden R, Vart P, Franssen CFM, Hemmelder MH, Jager KJ, et al. COVID-19-related mortality in kidney transplant and dialysis patients: results of the ERACODA collaboration. Nephrol Dial Transplant. 2020;35(11):1973–83. Epub 2020/11/06. doi: 10.1093/ndt/gfaa261. PubMed PMID: 33151337; PubMed Central PMCID: PMCPMC7665620.

9. The Renal Association B, UK. UK Renal Registry (2020) COVID-19 surveillance report for renal centres in the UK: All regions and centres.

10. Robinson BM, Guedes M, Alghonaim M, Cases A, Dasgupta I, Gan L, et al. Worldwide Early Impact of COVID-19 on Dialysis Patients and Staff and Lessons Learned: A DOPPS Roundtable Discussion. Kidney Med. 2021;3(4):619–34. Epub 2021/05/20. doi: 10.1016/j.xkme.2021.03.006. PubMed PMID: 34007963; PubMed Central PMCID: PMCPMC8120787.

11. Baden LR, El Sahly HM, Essink B, Kotloff K, Frey S, Novak R, et al. Efficacy and Safety of the mRNA-1273 SARS-CoV-2 Vaccine. N Engl J Med. 2021;384(5):403–16. Epub 2020/12/31. doi: 10.1056/NEJMoa2035389. PubMed PMID: 33378609; PubMed Central PMCID: PMCPMC7787219.

12. Polack FP, Thomas SJ, Kitchin N, Absalon J, Gurtman A, Lockhart S, et al. Safety and Efficacy of the BNT162b2 mRNA Covid-19 Vaccine. N Engl J Med. 2020;383(27):2603–15. Epub 2020/12/11. doi: 10.1056/NEJMoa2034577. PubMed PMID: 33301246; PubMed Central PMCID: PMCPMC7745181.

13. Dagan N, Barda N, Kepten E, Miron O, Perchik S, Katz MA, et al. BNT162b2 mRNA Covid-19 Vaccine in a Nationwide Mass Vaccination Setting. N Engl J Med. 2021;384(15):1412–23. Epub 2021/02/25. doi: 10.1056/NEJMoa2101765. PubMed PMID: 33626250; PubMed Central PMCID: PMCPMC7944975.

14. Butt AA, Omer SB, Yan P, Shaikh OS, Mayr FB. SARS-CoV-2 Vaccine Effectiveness in a High-Risk National Population in a Real-World Setting. Ann Intern Med. 2021. Epub 2021/07/20. doi: 10.7326/M21-1577. PubMed PMID: 34280332; PubMed Central PMCID: PMCPMC8381771.

15. Malard F, Gaugler B, Gozlan J, Bouquet L, Fofana D, Siblany L, et al. Weak immunogenicity of SARS-CoV-2 vaccine in patients with hematologic malignancies. Blood Cancer J. 2021;11(8):142. Epub 2021/08/12. doi: 10.1038/s41408-021-00534-z. PubMed PMID: 34376633; PubMed Central PMCID: PMCPMC8353615.

16. Louapre C, Ibrahim M, Maillart E, Abdi B, Papeix C, Stankoff B, et al. Anti-CD20 therapies decrease humoral immune response to SARS-CoV-2 in patients with multiple sclerosis or neuromyelitis optica spectrum disorders. J Neurol Neurosurg Psychiatry. 2021. Epub 2021/08/04. doi: 10.1136/jnnp-2021-326904. PubMed PMID: 34341142; PubMed Central PMCID: PMCPMC8331322.

17. Boyarsky BJ, Werbel WA, Avery RK, Tobian AAR, Massie AB, Segev DL, et al. Antibody Response to 2-Dose SARS-CoV-2 mRNA Vaccine Series in Solid Organ Transplant Recipients. JAMA. 2021;325(21):2204–6. Epub 2021/05/06. doi: 10.1001/jama.2021.7489. PubMed PMID: 33950155; PubMed Central PMCID: PMCPMC8100911.

18. Ikizler TA, Coates PT, Rovin BH, Ronco P. Immune response to SARS-CoV-2 infection and vaccination in patients receiving kidney replacement therapy. Kidney Int. 2021;99(6):1275–9. Epub 2021/05/02. doi: 10.1016/j.kint.2021.04.007. PubMed PMID: 33931226; PubMed Central PMCID: PMCPMC8055920.

19. Rincon-Arevalo H, Choi M, Stefanski AL, Halleck F, Weber U, Szelinski F, et al. Impaired humoral immunity to SARS-CoV-2 BNT162b2 vaccine in kidney transplant recipients and dialysis patients. Sci Immunol. 2021;6(60). Epub 2021/06/17. doi: 10.1126/sciimmunol.abj1031. PubMed PMID: 34131023.

20. Danthu C, Hantz S, Dahlem A, Duval M, Ba B, Guibbert M, et al. Humoral Response after SARS-CoV-2 mRNA Vaccination in a Cohort of Hemodialysis Patients and Kidney Transplant Recipients. J Am Soc Nephrol. 2021. Epub 2021/06/18. doi: 10.1681/ASN.2021040490. PubMed PMID: 34135083.

21. Public Health England, Immunization against infection disease: the Green book: Public Health England; 2021.

22. Grupper A, Sharon N, Finn T, Cohen R, Israel M, Agbaria A, et al. Humoral Response to the Pfizer BNT162b2 Vaccine in Patients Undergoing Maintenance Hemodialysis. Clin J Am Soc Nephrol. 2021. Epub 2021/04/08. doi: 10.2215/CJN.03500321. PubMed PMID: 33824157.

23. Zitt E, Davidovic T, Schimpf J, Abbassi-Nik A, Mutschlechner B, Ulmer H, et al. The Safety and Immunogenicity of the mRNA-BNT162b2 SARS-CoV-2 Vaccine in Hemodialysis Patients. Front Immunol. 2021;12:704773. Epub 2021/07/06. doi: 10.3389/fimmu.2021.704773. PubMed PMID: 34220867; PubMed Central PMCID: PMCPMC8242233.

24. Alcazar-Arroyo R, Portoles J, Lopez-Sanchez P, Zalamea F, Furaz K, Mendez A, et al. Rapid decline of anti-SARS-CoV-2 antibodies in patients on haemodialysis: the COVID-FRIAT study. Clin Kidney J. 2021;14(7):1835–44. Epub 2021/07/03. doi: 10.1093/ckj/sfab048. PubMed PMID: 34211708; PubMed Central PMCID: PMCPMC7989535.

25. Krueger KM, Ison MG, Ghossein C. Practical Guide to Vaccination in All Stages of CKD, Including Patients Treated by Dialysis or Kidney Transplantation. Am J Kidney Dis. 2020;75(3):417–25. Epub 2019/10/06. doi: 10.1053/j.ajkd.2019.06.014. PubMed PMID: 31585683.

26. Chaves SS, Daniels D, Cooper BW, Malo-Schlegel S, Macarthur S, Robbins KC, et al. Immunogenicity of hepatitis B vaccine among hemodialysis patients: effect of revaccination of non-responders and duration of protection. Vaccine. 2011;29(52):9618–23. Epub 2011/11/03. doi: 10.1016/j.vaccine.2011.10.057. PubMed PMID: 22044739.

27. Mlcochova P, Kemp S, Dhar MS, Papa G, Meng B, Ferreira I, et al. SARS-CoV-2 B.1.617.2 Delta variant replication and immune evasion. Nature. 2021. Epub 2021/09/07. doi: 10.1038/s41586-021-03944-y. PubMed PMID: 34488225.

28. Kirchdoerfer RN, Wang N, Pallesen J, Wrapp D, Turner HL, Cottrell CA, et al. Stabilized coronavirus spikes are resistant to conformational changes induced by receptor recognition or proteolysis. Sci Rep. 2018;8(1):15701. Epub 2018/10/26. doi: 10.1038/s41598-018-34171-7. PubMed PMID: 30356097; PubMed Central PMCID: PMCPMC6200764.

29. Pallesen J, Wang N, Corbett KS, Wrapp D, Kirchdoerfer RN, Turner HL, et al. Immunogenicity and structures of a rationally designed prefusion MERS-CoV spike antigen. Proc Natl Acad Sci U S A. 2017;114(35):E7348–E57. Epub 2017/08/16. doi: 10.1073/pnas.1707304114. PubMed PMID: 28807998; PubMed Central PMCID: PMCPMC5584442.

30. Walls AC, Park YJ, Tortorici MA, Wall A, McGuire AT, Veesler D. Structure, Function, and Antigenicity of the SARS-CoV-2 Spike Glycoprotein. Cell. 2020;181(2):281–92 e6. Epub 2020/03/11. doi: 10.1016/j.cell.2020.02.058. PubMed PMID: 32155444; PubMed Central PMCID: PMCPMC7102599.

31. Gregson J, Rhee SY, Datir R, Pillay D, Perno CF, Derache A, et al. Human Immunodeficiency Virus-1 Viral Load Is Elevated in Individuals With Reverse-Transcriptase Mutation M184V/I During Virological Failure of First-Line Antiretroviral Therapy and Is Associated With Compensatory Mutation L74I. J Infect Dis. 2020;222(7):1108–16. Epub 2019/11/28. doi: 10.1093/infdis/jiz631. PubMed PMID: 31774913; PubMed Central PMCID: PMCPMC7459140.

32. Forloni M, Liu AY, Wajapeyee N. Creating Insertions or Deletions Using Overlap Extension Polymerase Chain Reaction (PCR) Mutagenesis. Cold Spring Harb Protoc. 2018;2018(8). Epub 2018/08/03. doi: 10.1101/pdb.prot097758. PubMed PMID: 30068588.

33. Takada A, Robison C, Goto H, Sanchez A, Murti KG, Whitt MA, et al. A system for functional analysis of Ebola virus glycoprotein. Proc Natl Acad Sci U S A. 1997;94(26):14764–9. Epub 1998/02/07. doi: 10.1073/pnas.94.26.14764. PubMed PMID: 9405687; PubMed Central PMCID: PMCPMC25111.

34. Riblett AM, Blomen VA, Jae LT, Altamura LA, Doms RW, Brummelkamp TR, et al. A Haploid Genetic Screen Identifies Heparan Sulfate Proteoglycans Supporting Rift Valley Fever Virus Infection. J Virol. 2016;90(3):1414–23. Epub 2015/11/20. doi: 10.1128/JVI.02055-15. PubMed PMID: 26581979; PubMed Central PMCID: PMCPMC4719632.

35. McCallum M, Bassi J, De Marco A, Chen A, Walls AC, Di Iulio J, et al. SARS-CoV-2 immune evasion by the B.1.427/B.1.429 variant of concern. Science. 2021;373(6555):648–54. Epub 2021/07/03. doi: 10.1126/science.abi7994. PubMed PMID: 34210893.

36. Piccoli L, Park YJ, Tortorici MA, Czudnochowski N, Walls AC, Beltramello M, et al. Mapping Neutralizing and Immunodominant Sites on the SARS-CoV-2 Spike Receptor-Binding Domain by Structure-Guided High-Resolution Serology. Cell. 2020;183(4):1024–42 e21. Epub 2020/09/30. doi: 10.1016/j.cell.2020.09.037. PubMed PMID: 32991844; PubMed Central PMCID: PMCPMC7494283.

37. Legros V, Denolly S, Vogrig M, Boson B, Siret E, Rigaill J, et al. A longitudinal study of SARS-CoV-2-infected patients reveals a high correlation between neutralizing antibodies and COVID-19 severity. Cell Mol Immunol. 2021;18(2):318–27. Epub 2021/01/08. doi: 10.1038/s41423-020-00588-2. PubMed PMID: 33408342; PubMed Central PMCID: PMCPMC7786875.

38. Roltgen K, Powell AE, Wirz OF, Stevens BA, Hogan CA, Najeeb J, et al. Defining the features and duration of antibody responses to SARS-CoV-2 infection associated with disease severity and outcome. Sci Immunol. 2020;5(54). Epub 2020/12/09. doi: 10.1126/sciimmunol.abe0240. PubMed PMID: 33288645; PubMed Central PMCID: PMCPMC7857392.

39. Kliger AS. Targeting Zero Infections in Dialysis: New Devices, Yes, but also Guidelines, Checklists, and a Culture of Safety. J Am Soc Nephrol. 2018;29(4):1083–4. Epub 2018/03/07. doi: 10.1681/ASN.2018020132. PubMed PMID: 29507046; PubMed Central PMCID: PMCPMC5875966.

40. Windpessl M, Bruchfeld A, Anders HJ, Kramer H, Waldman M, Renia L, et al. COVID-19 vaccines and kidney disease. Nat Rev Nephrol. 2021;17(5):291–3. Epub 2021/02/10. doi: 10.1038/s41581-021-00406-6. PubMed PMID: 33558753; PubMed Central PMCID: PMCPMC7869766.

41. Schrezenmeier E, Bergfeld L, Hillus D, Lippert JD, Weber U, Tober-Lau P, et al. Immunogenicity of COVID-19 Tozinameran Vaccination in Patients on Chronic Dialysis. Front Immunol. 2021;12:690698. Epub 2021/07/20. doi: 10.3389/fimmu.2021.690698. PubMed PMID: 34276681; PubMed Central PMCID: PMCPMC8284337.

42. Stumpf J, Siepmann T, Lindner T, Karger C, Schwobel J, Anders L, et al. Humoral and cellular immunity to SARS-CoV-2 vaccination in renal transplant versus dialysis patients: A prospective, multicenter observational study using mRNA-1273 or BNT162b2 mRNA vaccine. Lancet Reg Health Eur. 2021:100178. Epub 2021/07/29. doi: 10.1016/j.lanepe.2021.100178. PubMed PMID: 34318288; PubMed Central PMCID: PMCPMC8299287.

43. Lacson E, Argyropoulos C, Manley H, Aweh G, Chin A, Salman L, et al. Immunogenicity of SARS-CoV-2 Vaccine in Dialysis. J Am Soc Nephrol. 2021. Epub 2021/08/06. doi: 10.1681/ASN.2021040432. PubMed PMID: 34348908.

44. Anderegg MA, Liu M, Saganas C, Montani M, Vogt B, Huynh-Do U, et al. De novo vasculitis after mRNA-1273 (Moderna) vaccination. Kidney Int. 2021;100(2):474–6. Epub 2021/06/05. doi: 10.1016/j.kint.2021.05.016. PubMed PMID: 34087251; PubMed Central PMCID: PMCPMC8166777.

45. Khoury DS, Cromer D, Reynaldi A, Schlub TE, Wheatley AK, Juno JA, et al. Neutralizing antibody levels are highly predictive of immune protection from symptomatic SARS-CoV-2 infection. Nat Med. 2021. Epub 2021/05/19. doi: 10.1038/s41591-021-01377-8. PubMed PMID: 34002089.

46. Speer C, Benning L, Tollner M, Nusshag C, Kalble F, Reichel P, et al. Neutralizing antibody response against variants of concern after vaccination of dialysis patients with BNT162b2. Kidney Int. 2021;100(3):700–2. Epub 2021/07/16. doi: 10.1016/j.kint.2021.07.002. PubMed PMID: 34265359; PubMed Central PMCID: PMCPMC8274271.

47. Cohn BA, Cirillo PM, Murphy CC, Krigbaum NY, Wallace AW. SARS-CoV-2 vaccine protection and deaths among US veterans during 2021. Science. 2021:eabm0620. Epub 20211104. doi: 10.1126/science.abm0620. PubMed PMID: 34735261.

48. Hazell SL. Clinical utility of avidity assays. Expert Opin Med Diagn. 2007;1(4):511–9. Epub 2007/12/01. doi: 10.1517/17530059.1.4.511. PubMed PMID: 23496357.

49. Gaspar EB, De Gaspari E. Avidity assay to test functionality of anti-SARS-Cov-2 antibodies. Vaccine. 2021;39(10):1473–5. Epub 2021/02/15. doi: 10.1016/j.vaccine.2021.02.003. PubMed PMID: 33581919; PubMed Central PMCID: PMCPMC7857056.

50. Bauer G, Struck F, Schreiner P, Staschik E, Soutschek E, Motz M. The challenge of avidity determination in SARS-CoV-2 serology. J Med Virol. 2021;93(5):3092–104. Epub 2021/02/11. doi: 10.1002/jmv.26863. PubMed PMID: 33565617; PubMed Central PMCID: PMCPMC8013859.

51. Teimouri A, Mohtasebi S, Kazemirad E, Keshavarz H. Role of Toxoplasma gondii IgG Avidity Testing in Discriminating between Acute and Chronic Toxoplasmosis in Pregnancy. J Clin Microbiol. 2020;58(9). Epub 2020/04/24. doi: 10.1128/JCM.00505-20. PubMed PMID: 32321784; PubMed Central PMCID: PMCPMC7448626.

52. Genco F, Sarasini A, Parea M, Prestia M, Scudeller L, Meroni V. Comparison of the LIAISON(R)XL and ARCHITECT IgG, IgM, and IgG avidity assays for the diagnosis of Toxoplasma, cytomegalovirus, and rubella virus infections. New Microbiol. 2019;42(2):88–93. Epub 2019/04/18. PubMed PMID: 30994178.

53. Chan L, Fuca N, Zeldis E, Campbell KN, Shaikh A. Antibody Response to mRNA-1273 SARS-CoV-2 Vaccine in Hemodialysis Patients with and without Prior COVID-19. Clin J Am Soc Nephrol. 2021;16(8):1258–60. Epub 2021/05/26. doi: 10.2215/CJN.04080321. PubMed PMID: 34031182.

54. Carr EJ, Wu M, Harvey R, Wall EC, Kelly G, Hussain S, et al. Neutralising antibodies after COVID-19 vaccination in UK haemodialysis patients. The Lancet. 2021. doi: 10.1016/s0140-6736(21)01854-7.

55. Chapin-Bardales J, Gee J, Myers T. Reactogenicity Following Receipt of mRNA-Based COVID-19 Vaccines. JAMA. 2021;325(21):2201–2. Epub 2021/04/06. doi: 10.1001/jama.2021.5374. PubMed PMID: 33818592.

56. Puranik A, Lenehan PJ, Silvert E, Niesen MJM, Corchado-Garcia J, O’Horo JC, et al. Comparison of two highly-effective mRNA vaccines for COVID-19 during periods of Alpha and Delta variant prevalence. medRxiv. 2021. Epub 2021/08/18. doi: 10.1101/2021.08.06.21261707. PubMed PMID: 34401884.

57. Abe KT, Hu Q, Mozafarihashjin M, Samson R, Manguiat K, Robinson A, et al. Neutralizing antibody responses to SARS-CoV-2 variants in vaccinated Ontario long-term care home residents and workers. medRxiv. 2021. doi: 10.1101/2021.08.06.21261721v2.

58. Steensels D, Pierlet N, Penders J, Mesotten D, Heylen L. Comparison of SARS-CoV-2 Antibody Response Following Vaccination With BNT162b2 and mRNA-1273. JAMA. 2021;326(15):1533–5. Epub 2021/08/31. doi: 10.1001/jama.2021.15125. PubMed PMID: 34459863; PubMed Central PMCID: PMCPMC8406205.

59. Richards NE, Keshavarz B, Workman LJ, Nelson MR, Platts-Mills TAE, Wilson JM. Comparison of SARS-CoV-2 Antibody Response by Age Among Recipients of the BNT162b2 vs the mRNA-1273 Vaccine. JAMA Netw Open. 2021;4(9):e2124331. Epub 2021/09/03. doi: 10.1001/jamanetworkopen.2021.24331. PubMed PMID: 34473262; PubMed Central PMCID: PMCPMC8414189.

60. Jahn M, Korth J, Dorsch O, Anastasiou OE, Sorge-Hadicke B, Tyczynski B, et al. Humoral Response to SARS-CoV-2-Vaccination with BNT162b2 (Pfizer-BioNTech) in Patients on Hemodialysis. Vaccines (Basel). 2021;9(4). Epub 2021/05/01. doi: 10.3390/vaccines9040360. PubMed PMID: 33918085; PubMed Central PMCID: PMCPMC8070660.

61. Clarke CL, Prendecki M, Dhutia A, Gan J, Edwards C, Prout V, et al. Longevity of SARS-CoV-2 immune responses in hemodialysis patients and protection against reinfection. Kidney Int. 2021;99(6):1470–7. Epub 2021/03/29. doi: 10.1016/j.kint.2021.03.009. PubMed PMID: 33774082; PubMed Central PMCID: PMCPMC7992297.

62. Longlune N, Nogier MB, Miedouge M, Gabilan C, Cartou C, Seigneuric B, et al. High immunogenicity of a messenger RNA based vaccine against SARS-CoV-2 in chronic dialysis patients. Nephrol Dial Transplant. 2021. Epub 2021/06/01. doi: 10.1093/ndt/gfab193. PubMed PMID: 34057463; PubMed Central PMCID: PMCPMC8195197.

63. Ducloux D, Colladant M, Chabannes M, Yannaraki M, Courivaud C. Humoral response after 3 doses of the BNT162b2 mRNA COVID-19 vaccine in patients on hemodialysis. Kidney Int. 2021;100(3):702–4. Epub 2021/07/04. doi: 10.1016/j.kint.2021.06.025. PubMed PMID: 34216675; PubMed Central PMCID: PMCPMC8243640.

64. Espi M, Charmetant X, Barba T, Mathieu C, Pelletier C, Koppe L, et al. A prospective observational study for justification, safety, and efficacy of a third dose of mRNA vaccine in patients receiving maintenance hemodialysis. Kidney Int. 2021. Epub 20211129. doi: 10.1016/j.kint.2021.10.040. PubMed PMID: 34856313; PubMed Central PMCID: PMCPMC8628628.

65. Dekervel M, Henry N, Torreggiani M, Pouteau LM, Imiela JP, Mellaza C, et al. Humoral response to a third injection of BNT162b2 vaccine in patients on maintenance haemodialysis. Clin Kidney J. 2021;14(11):2349–55. Epub 20210813. doi: 10.1093/ckj/sfab152. PubMed PMID: 34754430; PubMed Central PMCID: PMCPMC8573007.

66. Massa F, Cremoni M, Gerard A, Grabsi H, Rogier L, Blois M, et al. Safety and cross-variant immunogenicity of a three-dose COVID-19 mRNA vaccine regimen in kidney transplant recipients. EBioMedicine. 2021;73:103679. Epub 20211108. doi: 10.1016/j.ebiom.2021.103679. PubMed PMID: 34763205; PubMed Central PMCID: PMCPMC8573385.

67. Robert T, Lano G, Giot M, Fourie T, de Lamballeri X, Jehel O, et al. Humoral response after SARS-COV2 vaccination in patient undergoing maintenance hemodialysis: loss of immunity, third dose and non-responders. Nephrol Dial Transplant. 2021. Epub 20211013. doi: 10.1093/ndt/gfab299. PubMed PMID: 34643714.

