## Supplementary Information for "Poor neutralization and rapid decay of antibodies to SARS-CoV-2 variants in vaccinated dialysis patients"

### Supporting Information

#### S1 Fig

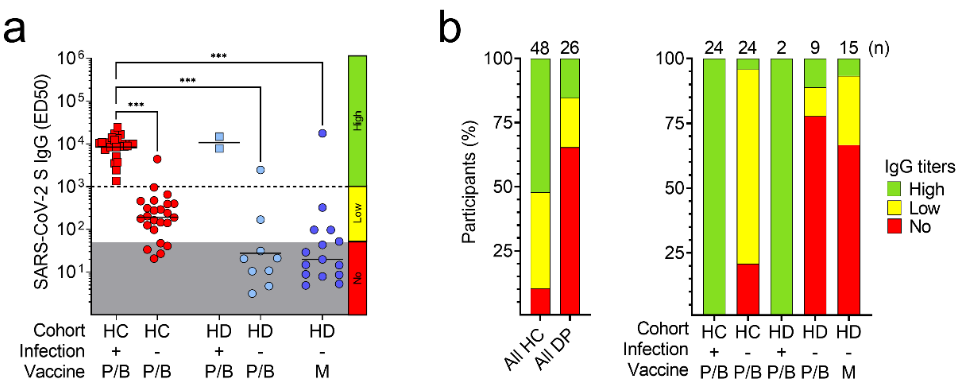


**S1 Fig. Plasma antibody titers to SARS-CoV-2 after one dose of mRNA vaccine.** a) Plasma IgG titers (ED50) to SARS-CoV-2 S after one dose of Pfizer/BioNTech (P/B) or Moderna (M) vaccines in previously infected (square) and naïve (circle) healthy controls (HC, red) and hemodialysis (HD, blue) patients. Grey areas indicate non-specific IgG titers <50, a cut-off that was determined on non-specific binding to uncoated ELISA plates. An additional cut-off of 1’000, determined from the lowest titers in previously infected HC, was used to distinguish low (50-1’000) from high (>1’000) IgG titers. Statistical significance is set as P<0.05 and P-values are indicated with asterisks (*=0.033; **=0.002; ***<0.001). b) Percentages of participants with high, low or no plasma IgG to SARS-CoV-2 S after one dose of mRNA-vaccine. Total number of participants within each cohort of HC and dialysis patients (DP) and within each subgroup is shown at the top of each bar.

### S2 Fig

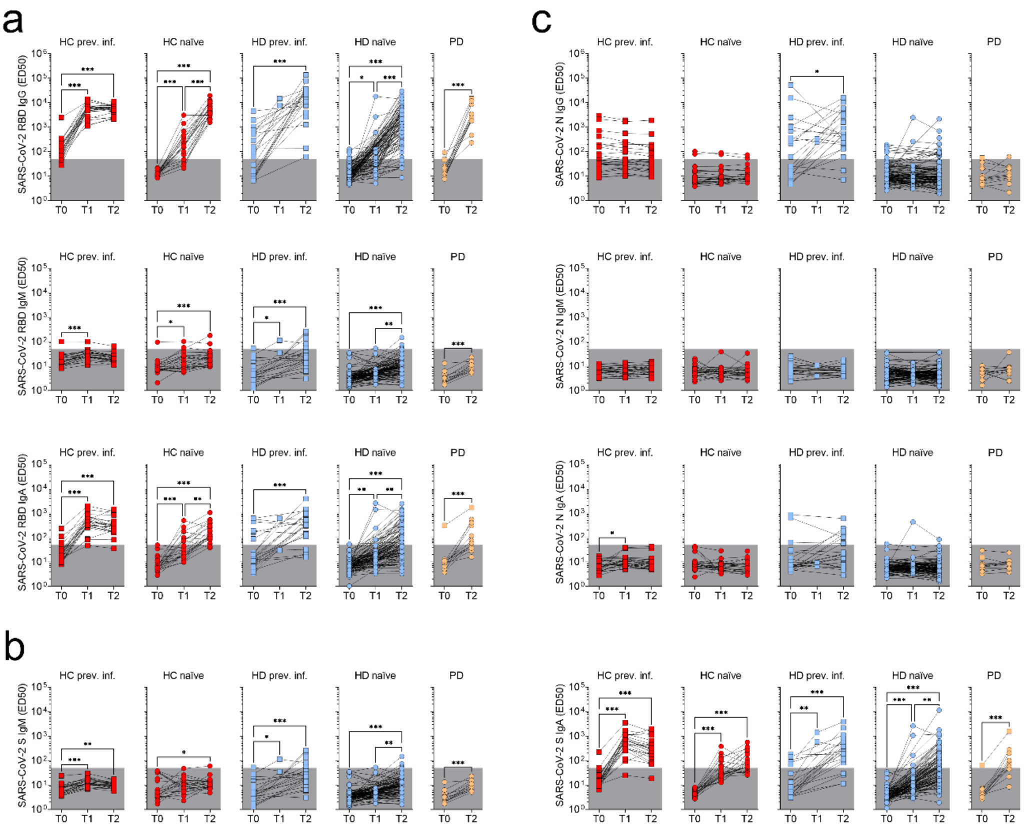


**S2 Fig. Kinetics of plasma antibody titers to SARS-CoV-2 in healthy controls and dialysis patients.** a) Plasma IgG, IgM and IgA titers to SARS-CoV-2 RBD. b) Plasma IgM and IgA titers (ED50) to SARS-CoV-2 S measured before vaccination (T0), after one (T1) or two (T2) vaccine doses. c) Plasma IgG, IgM and IgA titers to SARS-CoV-2 Nucleoprotein (N).

### S3 Fig

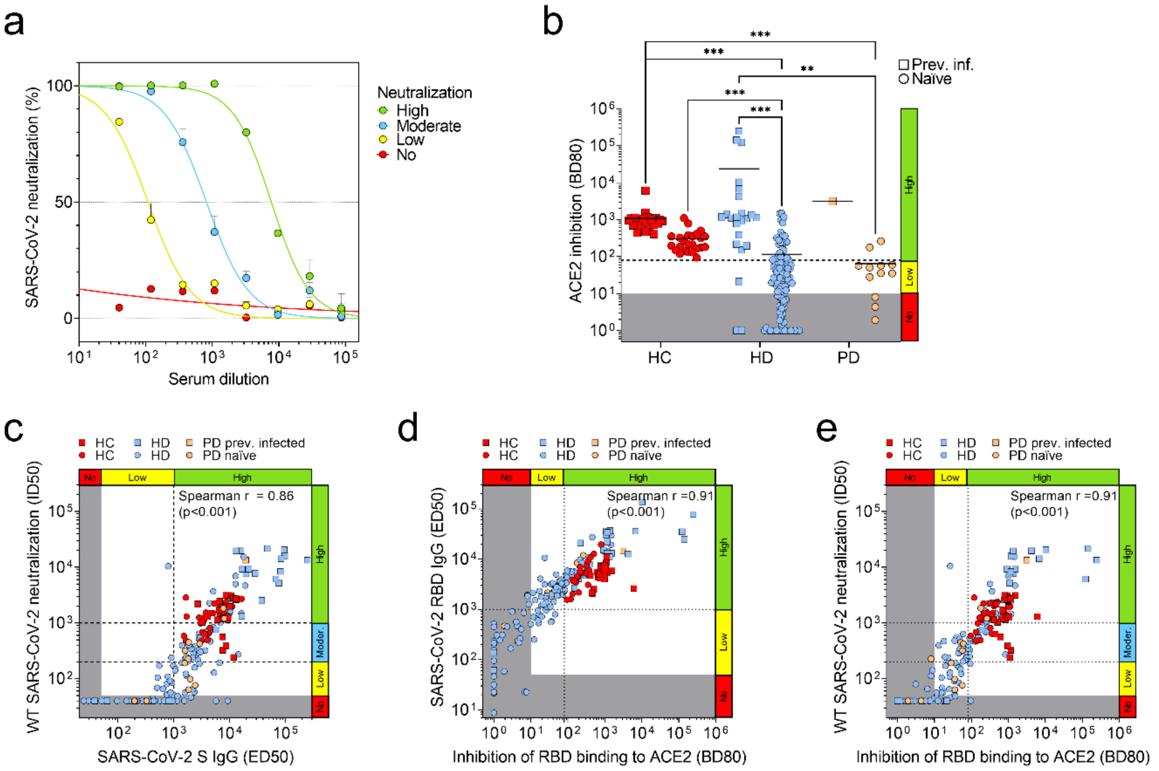


**S3 Fig. Correlation of plasma IgG titers, ACE2 inhibition and neutralization against wild-type SARS-CoV-2.** a) Neutralization of WT SARS-CoV-2 pseudotyped VSV by four representative plasma samples showing high, moderate, low or no neutralization. b) Inhibition of RBD binding to human ACE2 by plasma antibodies in previously infected (square) and naïve (circle) healthy controls (HC), hemodialysis (HD) and peritoneal dialysis (PD) patients after two doses of mRNA vaccine. Grey areas indicate no ACE2 inhibition by plasma antibodies with BD80 titers lower than 10. A cut-off of 80, determined from the lowest inhibition capacity in HC, was used to distinguish low (10-80) from high (>80) inhibition. Statistical significance is set as P<0.05 and P-values are indicated with asterisks (*=0.033; **=0.002; ***<0.001). Shown are data from n=2 independent experiments. c) Correlation analysis between plasma IgG titers index and WT SARS-CoV-2 neutralization in all the plasma samples collected after the second vaccine dose. d) Correlation analysis between ACE2 inhibition and plasma RBD IgG titers index in all the plasma samples collected after the second vaccine dose. e) Correlation analysis between ACE2 inhibition and WT SARS-CoV-2 neutralization in all the plasma samples collected after the second vaccine dose.

### S4 Fig

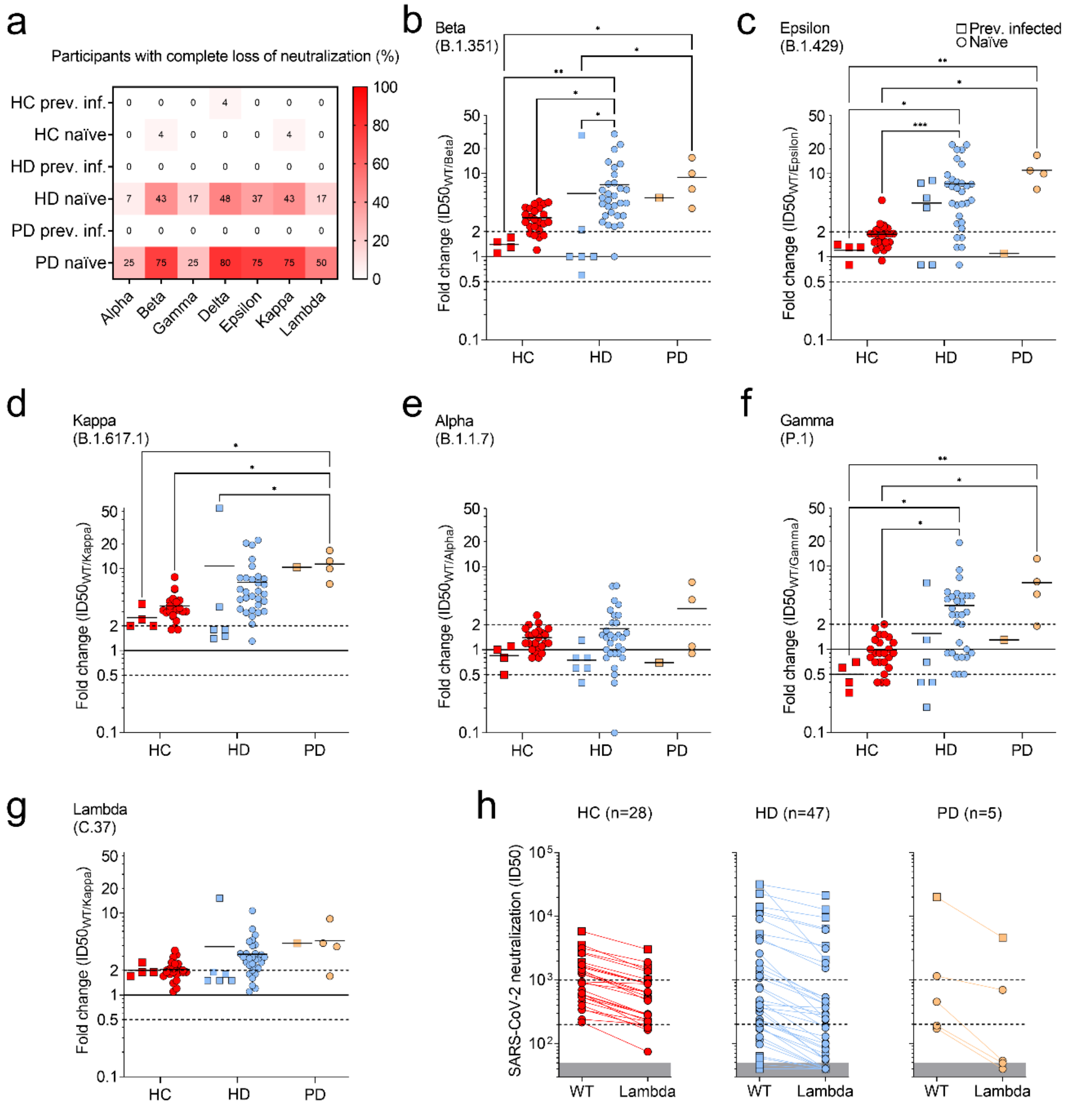


**S4 Fig. Comparison of neutralizing antibody titers against wild-type SARS-CoV-2 and other variants.** a) Fraction of participants with neutralizing titers against WT SARS-CoV-2 (ID50>40) showing complete loss of neutralization against Alpha (B.1.1.7), Beta (B.1.351), Gamma (P.1), Delta (B.1.617.2), Epsilon (B.1.429), Kappa (B.1.617.1) and Lambda (C.37) variants. b-g) Fold change analysis of neutralizing titers against WT and Beta (b), Epsilon (c), Kappa (d), Alpha (e), Gamma (f) and Lambda (g) SARS-CoV-2 in 28 HC, 36 HD and 5 PD patients with ID50 neutralizing titers against WT SARS-CoV-2 greater than 80. h) Side-by-side comparison of neutralizing titers against WT and Lambda (C.37) SARS-CoV-2 variant in 28 HC, 47 HD and 5 PD patients.

### S5 Fig

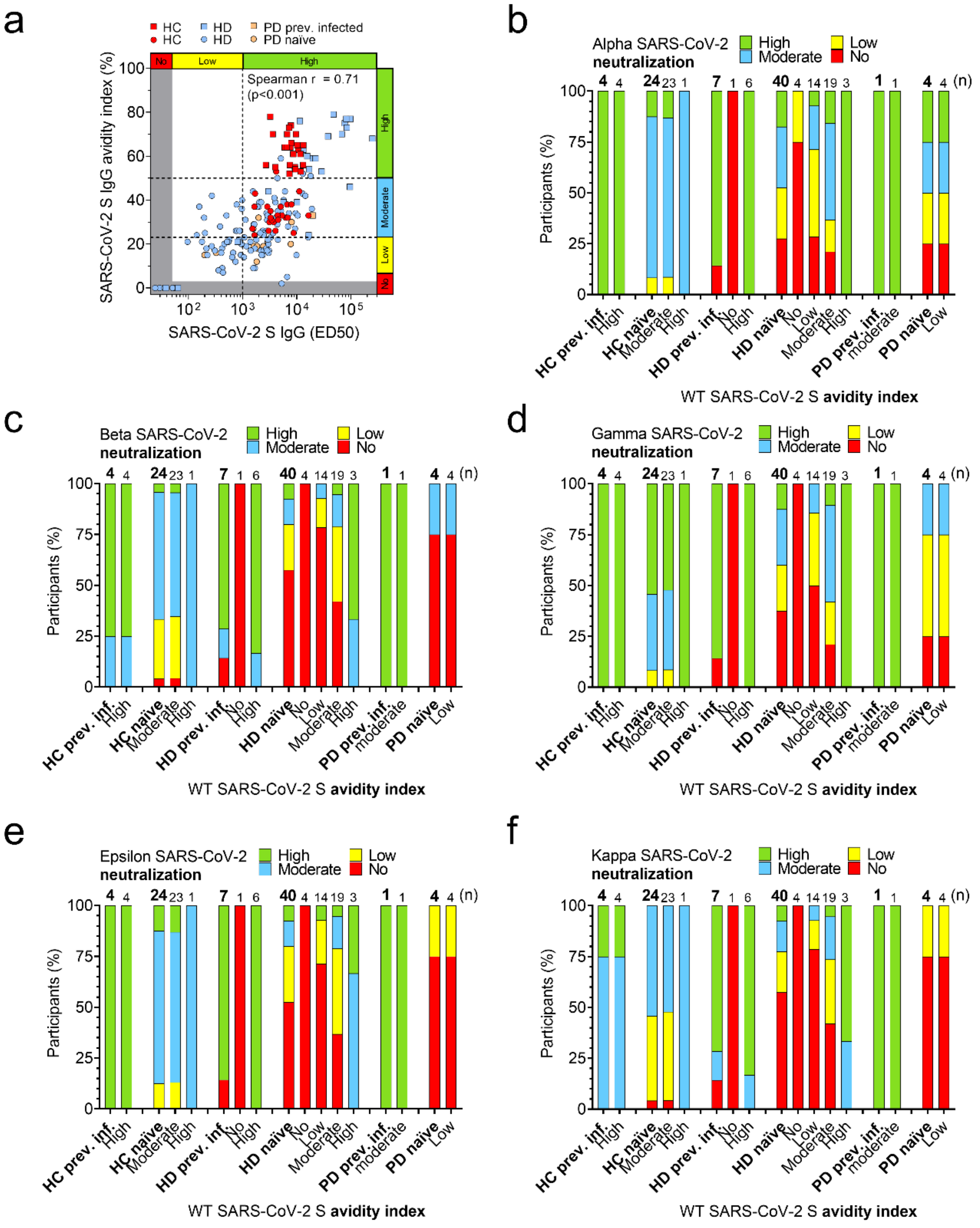


**S5 Fig. Correlation of avidity and neutralization of plasma antibodies to wild-type SARS-CoV-2 and other variants.** a) Correlation analysis between plasma IgG titers index and SARS-CoV-2 S IgG avidity in all the plasma samples collected after the second vaccine dose. b-f) Percentages of participants having plasma antibodies with high, moderate, low or no neutralization of Alpha (b), Beta (c), Gamma (d), Epsilon (e) and Kappa (f) SARS-CoV-2 after two doses of mRNA vaccine. Participants are shown as a total (bold) or divided by level of avidity to WT SARS-CoV-2 S (no, low, moderate, high). Total number of participants within each group is shown at the top of each bar.

#### S6 Fig

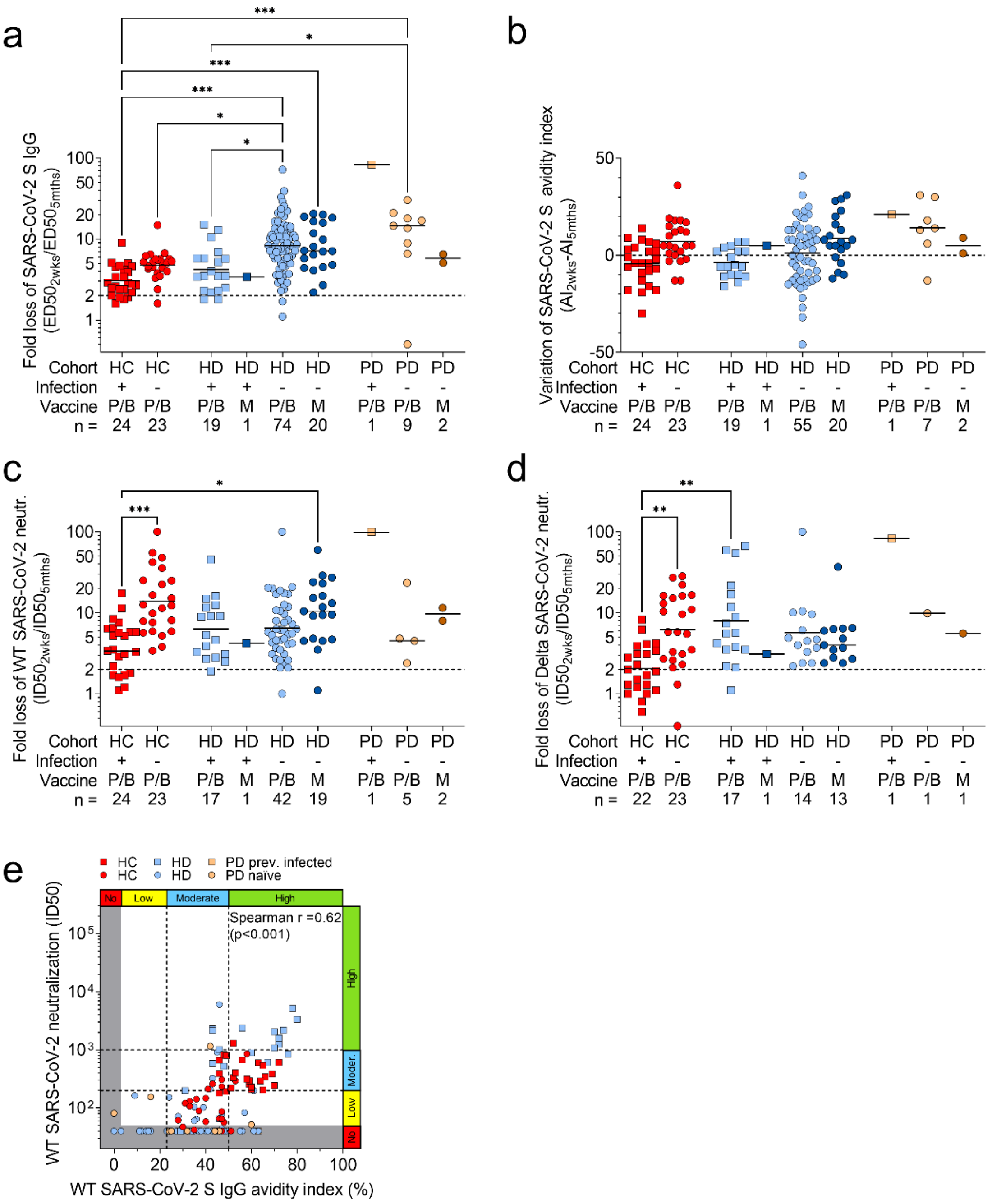


**S6 Fig. Comparison of variation of SARS-CoV-2 S IgG, neutralizing titers and avidity index.** a-d) Fold change analysis of SARS-CoV-2 S IgG (a), avidity index (b), neutralizing titers against WT SARS-CoV-2 (c) and Delta variant (d) measured in samples collected at 2 weeks and up to 7 months (average 5 months) after the second vaccine dose. n indicates the number of analyzed participants as explained in Table 4. e) Correlation analysis between plasma IgG avidity index to WT SARS-CoV-2 S and neutralization of WT SARS-CoV-2 in all the 180 plasma samples collected up to 7 months after the second vaccine dose.
